# An optimised serological machine learning model enabling targeted test-and-treat for *Plasmodium vivax* malaria

**DOI:** 10.64898/2026.08.09.26355326

**Authors:** Lauren Smith, Dionne C. Argyropoulos, Alison Paolo N. Bareng, Janise Lin, Nicholas Kiernan-Walker, Macie Lamont, Anju Abraham, Pailene S. Lim, Kenneth W. Wu, Timothy William, Nicholas M. Anstey, Matthew J. Grigg, Jetsumon Sattabongkot, Marcus Lacerda, Ventis Vahi, Ramin Mazhari, Ivo Mueller, Rhea J. Longley

## Abstract

The persistence of *Plasmodium vivax* is driven by the hidden reservoirs of infection, presenting a key obstacle to elimination. Antibodies persist after asexual infections are cleared from peripheral blood and therefore can indicate current and recent past infections. Here, we present a machine learning algorithm that classifies recent *P. vivax* infections using serological markers to identify likely hypnozoite carriers. Using serological measurements from year-long observational cohort studies conducted in three low-transmission settings (including negative controls, *N*=2,635), we selected optimal subsets of markers by balancing sero-diagnostic performance against assay complexity and scalability. We initially trained a random forest classifier and then subsequently we compared several machine learning classifiers. Tree-based methods consistently performed best, although differences were marginal. An online R Shiny application (PvSeroApp) was developed to automate data processing, quality control, and serostatus classification. This algorithm underpins the *P. vivax* serological testing and treatment (PvSeroTAT) strategy, enabling targeted anti-hypnozoite therapy and strengthening elimination efforts.

## 2. Introduction

Malaria remains a major global public health challenge, with the Global Technical Strategy target of reducing incidence by at least 90% by 2030 unlikely to be achieved^1,2^. Yet substantial progress has been made in lower-transmission settings in the Asia-Pacific and the Americas, with six countries certified as malaria free and a further seven countries having already achieved the 2030 targets. Unfortunately, recent global funding constraints and regional case fluctuations have slowed momentum towards regional elimination^1,3^. In these near- and pre-elimination settings, the proportion of malaria infections due to *Plasmodium vivax* is increasing^2,4^. The ability of *P. vivax* to form dormant liver-stage hypnozoites capable of reactivating long after the initial infection presents a key barrier to malaria elimination^5^. Current routine detection methods, including rapid diagnostic tests, microscopy and PCR, cannot detect these hypnozoites. Moreover, with *P. vivax* asexual biomass concentrated as hidden reservoirs in the spleen and bone marrow^5,6^, peripheral blood parasite densities are often very low, thus frequently escaping detection by routine diagnostic methods. Innovative diagnostics, interventions and tools are urgently needed to target these hidden reservoirs and accelerate elimination progress.

Radical cure is the cornerstone of *P. vivax* elimination strategies as it combines blood-stage treatment (chloroquine or artemisinin-based combination therapy) and liver-stage treatment (8-aminoquinoline, such as primaquine or tafenoquine, which also have synergistic activity against asexual stages). The key challenge lies in identifying who to safely and effectively treat. Individuals with glucose-6-dehydrogenase (G6PD) deficiency, an X-linked disorder affecting approximately 5-30% of people in malaria endemic regions, are at risk of severe haemolysis when treated with 8-aminoquinolines^7–10^. This poses challenges for strategies to rapidly reduce *P. vivax* transmission. For example, mass drug administration, which treats entire populations irrespective of infection status, is not currently recommended by the World Health Organisation (WHO). Mass screening and treatment offers a more targeted approach by treating only those with detectable blood-stage infections identified by RDT or microscopy; however, this strategy overlooks the substantial reservoirs of low-density peripheral blood-stage, the hidden asexual-stage biomass in spleen and bone-marrow, and dormant liver-stage infections^11^, underestimating the true burden of infection.

The *P. vivax* serology test and treat strategy (PvSeroTAT) addresses limitations of existing malaria elimination strategies^12^. This strategy is based on the principle that blood-stage *P. vivax* infections elicit robust, long-lasting IgG antibody responses to multiple *P. vivax* antigens^13^, even following low-density infections^14^. As antibodies persist after parasite clearance from the peripheral blood, they can serve as markers of recent and past exposure as well as of ongoing hidden asexual stage-infections no longer detectable in peripheral blood. While hypnozoites cannot be directly detected with current technology, all tropical and sub-tropical *P. vivax* strains typically relapse within six to nine months without liver-stage treatment^12,15^, therefore individuals infected within this window, who have not received treatment, are likely hypnozoite carriers. PvSeroTAT uses a panel of antibody markers that reflect exposure within this period to pinpoint those most at risk and target them for anti-hypnozoite 8-aminoquinoline therapy following G6PD testing. This therapy also has the added advantage of activity against concurrent otherwise-undetectable hidden asexual-stage reservoirs of infection^5^. Risk-based modelling indicates that PvSeroTAT can achieve higher treatment coverage and greater reductions in *P. vivax* prevalence than mass screening and treatment ^12^, and nearly matches the impact of mass drug administration while treating three-fold fewer individuals at an 80% sensitivity/specificity threshold^16^. At 80% radical cure coverage, a single round of PvSeroTAT is estimated to reduce prevalence by ∼25% across low-, moderate- and high-transmission settings. With four rounds delivered six months apart, the impact increases substantially, with reductions up to 75% in low-transmission settings and 40-50% in moderate-to-high transmission settings^17^. Furthermore, field studies show that PvSeroTAT is operationally feasible even in highly mobile malaria-endemic communities despite added logistical complexity of laboratory-based testing^18^.

The current version of PvSeroTAT employs a laboratory-based multiplex Luminex bead-based assay to quantify antigen-specific IgG responses in human sera, which are subsequently analysed using a supervised machine learning algorithm to classify serostatus^12^. In the definitive study, a panel of eight *P. vivax* serological exposure markers (PvSEM) were selected from over 300 candidates and used to train a random forest classifier on year-long longitudinal cohort data from Brazil, Thailand and the Solomon Islands, as well as negative controls from within country and Australia. This assay achieved approximately 80% sensitivity and specificity in distinguishing individuals who had a recent *P. vivax* infection (≤ 9 months) from individuals with older *P. vivax* infections (> 9 months) or no infections^12^. However, practical constraints in antigen production and suitability necessitate refinement of this panel. In this paper, we present a revised PvSEM panel that excludes PvMSP3 due to its large size, instability and high genetic complexity^19,20^, and PVX_112670 due to expression difficulties. We evaluate a panel of candidate antigens, identify the eight best performers, and reparameterise the PvSEM algorithm with this refined panel.

While random forests have been widely applied to sero-surveillance data for both *P. vivax* and *P. falciparum*^12,21^, other machine learning algorithms, including logistic regression, k-nearest neighbours, extreme gradient boosting (XGBoost), and support vector machines, have also been successfully used for serostatus classification in infectious diseases such as sexually transmitted infections^22,23^, influenza^24^ and leishmaniasis^25^. Multi-algorithm ensemble approaches have further enhanced the predictive performance of infection status from serology data in *Plasmodium* species^26–29^, underscoring the growing potential of integrating serology with advanced computational methods to improve malaria sero-surveillance. These collective advancements have prompted updates to both the original *P. vivax* eight-antigen panel and the underlying machine learning algorithm.

This study presents an evaluation of supervised machine learning algorithms to classify *P. vivax* serostatus based on year-long cohort studies in low-transmission settings in Thailand, Brazil, and the Solomon Islands and negative controls from various blood donor sources in Australia, Thailand and Brazil. Leveraging our training dataset of total IgG antibody responses, we aimed to (i) identify a top combination of serological markers to predict past and current *P. vivax* infection that balances performance against assay complexity and scalability, (ii) train and optimise hyperparameters of a random forest algorithm on the longitudinal cohort dataset, (iii) compare the random forest with other widely used machine learning classification algorithms to evaluate the top performing algorithm for this dataset, (iv) adjust the model for potential cross-reactive serological exposure markers, and (v) develop a graphical user-interface (GUI) to that enables assay quality control and easy-access to apply the model for varied users with differing levels of expertise in biostatistics.

## 3 Result

### 3.1 Anti-*P. vivax* Antibody Responses Across Cohorts in Training Dataset

IgG antibody responses to the *P. vivax* serological exposure markers were validated using plasma samples from three year-long geographically diverse observational cohorts in Thailand, Brazil and the Solomon Islands with a known history of *Plasmodium* infections in the preceding 12 months. Individuals in these cohorts were assessed monthly for *Plasmodium* spp. by quantitative PCR (qPCR) with continuous concurrent passive case-detection at local malaria clinics and hospitals. Plasma samples from the last visit of these cohorts (*n*=680, 886, and 709 in Thailand, Brazil and the Solomon Islands, respectively) (**Table 1**) were used to measure IgG responses in a multiplex Luminex assay to ten *P. vivax* antigens (**Table 2**) selected from an overall panel of fourteen antigens (**Figure 1A**), including the top eight antigens as defined previously^12^. 5.67% (129/2,275) of individuals in the cohorts had a concurrent *P. vivax* infection, detected by qPCR, at the time the plasma was collected (**Table 2**). Pairwise Pearson correlations showed statistically significant but weak-to-moderate relationships in relative antibody unit (RAU) responses between *P. vivax* antigens, suggesting that the top ten antigens capture complementary (non-redundant) information (**Figure 1B**). Overall, there was a pattern of decreasing IgG magnitude with increasing time since last *P. vivax* infection, with minimal reactivity in malaria-naïve negative control individuals from Bangkok, Rio de Janeiro and Melbourne for most antigens (**Figure 1C**). The exception was for Pvs16, a peptide at 91% purity, which showed elevated reactivity in malaria-naïve, negative control individuals.

**Table 1.** Demographic characteristics of the study sites and participants included in the training dataset. Year-long observational cohort studies were conducted in Kanchanaburi and Ratchaburi provinces (Thailand), Manaus (Brazil), and Ngella (Solomon Islands). *P. vivax* negative controls were from the Brazil Blood Donor Registry (Br Neg), Thai Red Cross (ThRC), Australian Red Cross (ARC), and the volunteer biospecimen donor registry (VBDR) in Victoria, Australia. Age was not provided from the Thai Red Cross or Brazil Blood Donor Registry.

| Characteristic | Year-long longitudinal cohorts |  |  | Negative controls |  |  |  | Total |
| --- | --- | --- | --- | --- | --- | --- | --- | --- |
|  | Thailand | Solomon Islands | Brazil | ThRC | Br Neg | ARC | VBDR |  |
| No. of participants<sup>a</sup> | 680 | 709 | 886 | 69 | 96 | 97 | 98 | 2,635 |
| Median age<sup>b</sup> | 23[9-41] | 6[3-9] | 25[10-47] | - | - | 52[38-59] | 39[29-51] | 12[6-38] |
| Female sex<sup>a</sup> | 367 (54.0) | 336 (47.4) | 450 (50.8) | - | - | 52 (53.6) | 67 (68.4) | 1,272 |
| Time since last PCR positive sample<sup>a,d</sup> |  |  |  |  |  |  |  |  |
| Current<sup>e</sup> | 15 | 37 | 77 | 0 | 0 | 0 | 0 | 129 |
| < 9 months | 22 | 159 | 142 | 0 | 0 | 0 | 0 | 323 |
| 9-12 months | 16 | 30 | 26 | 0 | 0 | 0 | 0 | 72 |
| Not detected | 627 | 660 | 464 |  |  |  |  | 1,751 |
| Never | 0 | 0 | 0 | 69 | 96 | 97 | 98 | 360 |

<sup>a</sup> Data reflect No. (% [n/N]) of participants sampled.
<sup>b</sup> Data reflect the median age (value [IQR]).
<sup>d</sup> Only participants who had at least one *P. vivax* infection recorded in the previous year were included.
<sup>e</sup> Defined as participants who were *P. vivax* PCR positive at the time of the final visit when plasma was collected.

**Table 2.** Top ten *P. vivax* proteins selected as our serological exposure markers.

| Gene ID (PlasmoDB) | Chr | Gene Annotation | Allele | Construct size (AA) | Expression system | Provider | Function |
| --- | --- | --- | --- | --- | --- | --- | --- |
| PVX_087885 | 1 | Rhoptry associated membrane antigen (**PvRAMA**) | Sal-1 | 634-709 | *E. coli* | ZiP Diagnostics | Blood-stage; Part of the GPI-anchored proteins on the merozoite.<sup>30</sup> |
| PVX_096995 | 2 | Tryptophan-rich Antigen (**Pv-fam-a**) | Sal-1 | 56-480 | *E. coli* | ZiP Diagnostics | Blood-stage; Unspecified.<sup>31</sup> |
| PVX_000930 | 3 | Sexual stage antigen 16 (**Pvs16**) | Sal-1 | 30-92 | Peptide | Genscript | Sexual-stage; Immature gametocyte marker.<sup>32</sup> |
| PVX_003770 | 4 | Merozoite Protein 5 (**PvMSP5**) | Sal-1 | 25-364 | *E. coli* | ZiP Diagnostics | Blood-stage; Part of the GPI anchored proteins on the merozoite.<sup>33</sup> |
| PVX_099980 | 7 | Merozoite protein 1, C-terminal 19kD fragment (**PvMSP1-19**) | Sal-1 | 1623-1715 | *E. coli* | ZiP Diagnostics | Blood-stage; Carried to newly invaded red blood cells following merozoite egress, remains on merozoite surface.<sup>34</sup> |
| PVX_094255 | 8 | Reticulocyte binding protein 2b (**PvRBP2b**) | Sal-1 | 169-470 | *E. coli* | Tham Laboratory, WEHI | Blood-stage; Binds transferrin receptor 1 to mediate recognition of reticulocytes. Essential for invasion.<sup>35,36</sup> |
| PVX_097625 | 10 | Merozoite Protein 8 (**PvMSP8**) | Sal-1 | 24-465 | Insect cells | WEHI Protein Production Facility | Blood-stage; Part of the GPI anchored proteins on the merozoite.<sup>37</sup> |
| PVX_084720 | 13 | Translocon of Exported Proteins (**PvPTEX150**) | Sal-1 | 23-908 | Insect cells | WEHI Protein Production Facility | Blood-stage; Unspecified in *P. vivax* but ortholog in *P. falciparum* is on parasitophorous vacuole membrane responsible for protein export to erythrocyte cytosol.<sup>38</sup> |
| PVX_086200 | 13 | Cysteine-rich small secreted protein (**PvCSS**) | Sal-1 | 22-381 | Mammalian | WEHI Protein Production Facility | Blood-stage; Ortholog in *P. knowlesi* and *P. falciparum* is essential for merozoite invasion. In *P. falciparum*, it is part of the essential PCRCR complex. It forms part of the *P. vivax* PTRAMP-CSS-Ripr complex.<sup>39</sup> |
| KMZ83376.1<sup>a</sup> | - | Erythrocyte binding-like protein II (**PvEBPII**) | Brazil I | 1-716 | Insect cells | WEHI Protein Production Facility | Blood stage; Erythrocyte invasion.<sup>40</sup> |

Abbreviations: Chr = Chromosome, AA = Amino Acid, GPI = Glycophosphatidylinositol
<sup>a</sup> GeneBank ID.

**Figure 1.**
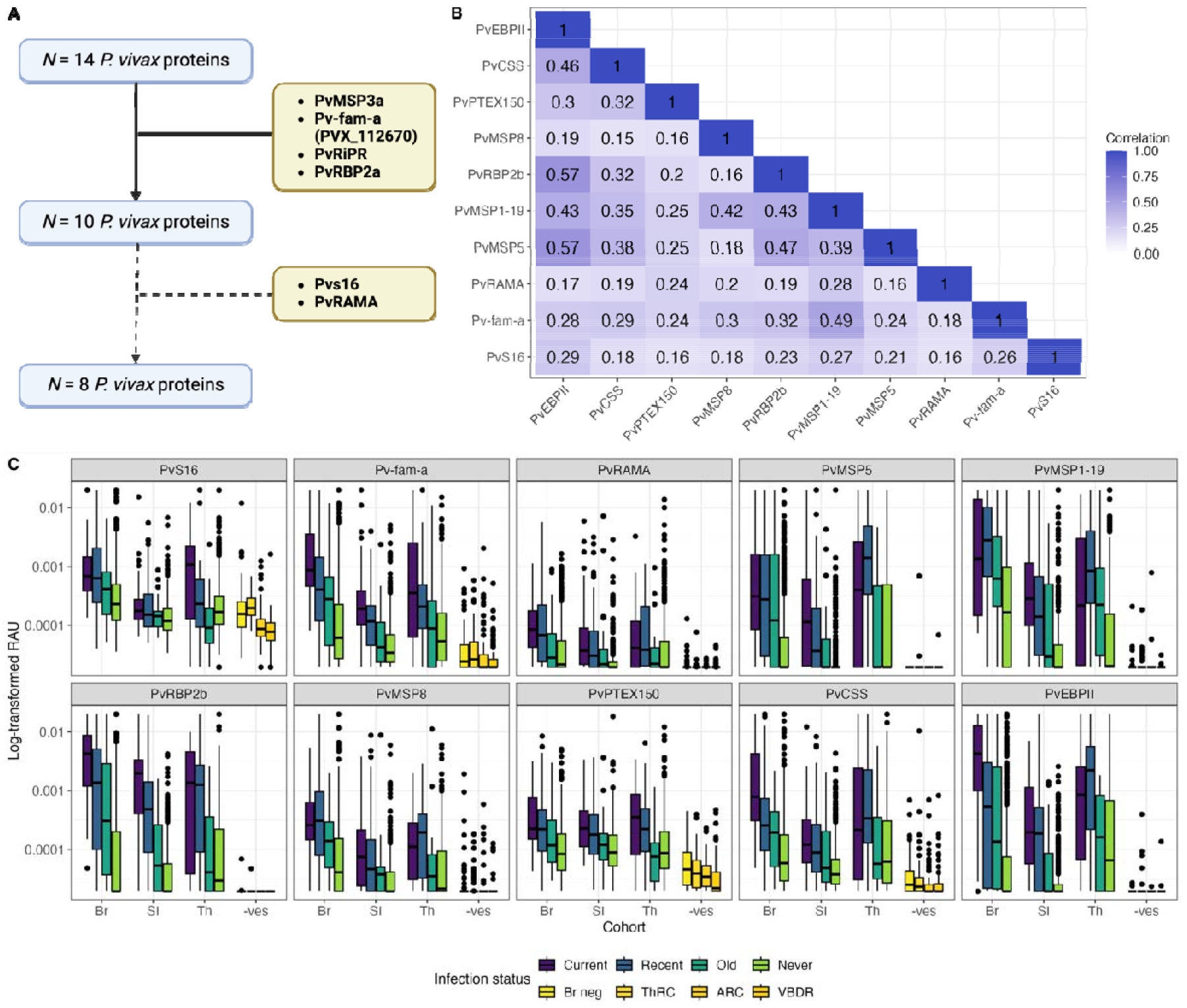
Selection of *P. vivax* serological exposure markers and IgG antibody responses considered in our panel. **(A)** Down-selection of *P. vivax* proteins considered to be serological exposure markers. Proteins not detailed in Table 2 are: *P. vivax* merozoite surface protein 3 (PvMSP3a, PVX_097720, chromosome 10, Sal-1), *P. vivax* tryptophan-rich antigen (Pv-fam-a, PVX_112670, unassigned, Sal-1), Rh5-interacting protein (PvRipr, PVP01_0816800, chromosome 8, P01) and reticulocyte binding protein 2a (PvRBP2a, PVP01_1402400, chromosome 14, P01). (**B**) Pairwise Pearson correlations between relative antibody units (RAU) for the top ten *P. vivax* antigens, with correlation coefficients ranging from 1 (dark blue) to 0 (white). All pairwise correlations were statistically significant. (**C**) IgG antibody responses in RAU for each *P. vivax* antigen considered in our panel from the year-long cohort studies in malaria endemic regions of Thailand (Th), the Solomon Islands (SI), and Brazil (Br), as well as negative controls from Brazil Blood Donor Registry (Br neg), Thai Red Cross (TRC), Australian Red Cross (ARC), the Australian Volunteer Biospecimen Donor Registry (VBDR). Infections from the year-long cohort studies were categorised as current (*P. vivax* infected at the final study time point, *N*=129), recent (*P. vivax* infected within the last 9 months, *N*=323), old (*P. vivax* infected 9-12 months, *N*=72) and never (never *P. vivax* infected during the study, *N*=1,751). **Table 1** provides a detailed breakdown of the *P. vivax* infection categories per cohort. Boxplots illustrate the median and 25th and 75th percentiles.

### 3.2 Evaluating Single and Multiple Antigen Classification Performance

We first evaluated the classification performance of each *P. vivax* antigen individually by calculating sensitivity and specificity across 1,000 cut-off thresholds and generating receiver operator characteristic (ROC) curves to assess their ability to distinguish recent from non-recent *P. vivax* infections. PvRBP2b had the highest area under the curve (AUC) value of 0.832 (standard error (SE): 0.012) providing the best single-antigen indicator of *P. vivax* exposure in the previous nine months (**Figure 2A**). PvRAMA and Pvs16 had AUC values less than 0.70 and were removed from further analysis (**Figure 1A, Figure 2A, Table S1**). Whilst Pvs16 had high background, background was low for PvRAMA and poorer performance was likely due to limited reactivity in individuals with current and recent *P. vivax* infection (**Figure 1C**).

**Figure 2.**
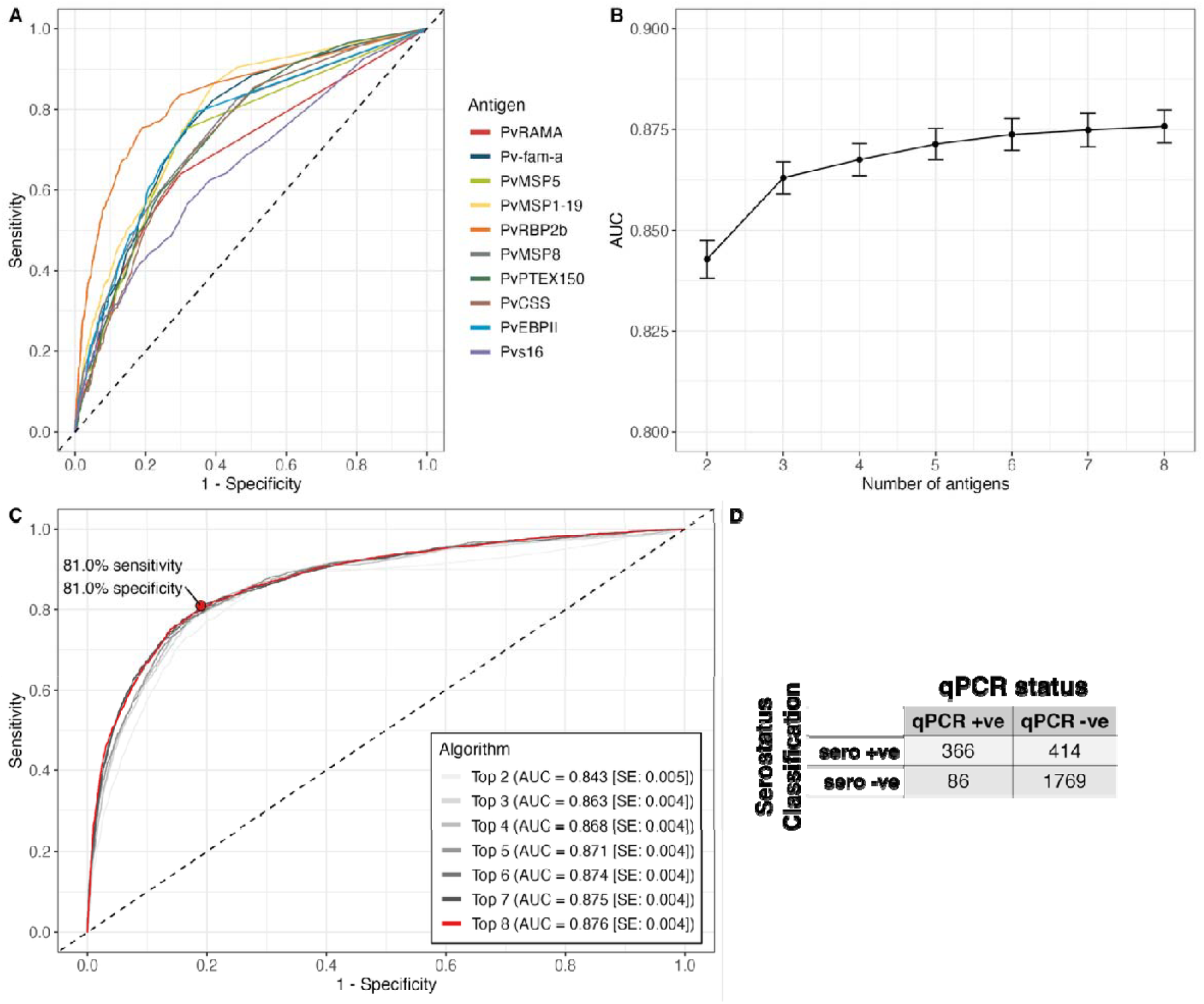
Classification performance of individual and multi-antigen *P. vivax* serological panels. **(A)** ROC curves for each of the ten individual *P. vivax* serological exposure markers (**B**) AUC values (with standard error) for the top-performing *P. vivax* antigen combinations, ranging from two to eight markers (Table 3). (**C**) ROC curves for the top-performing *P. vivax* multi-antigen combinations to illustrate the corresponding sensitivity and specificity profiles. The top 8 combination is outlined in red. (**D**) The confusion matrix provides a measure of the classifier’s accuracy and is a 2×2 table of the “Truth” defined from the *P. vivax* qPCR status from our longitudinal dataset (columns) and the models’ *P. vivax* serostatus “Prediction” when we classify these data (rows) based on our trained model. The Top Left are the **True Positives**, and the Bottom Right are the **True Negatives**.

To examine how *P. vivax* antigen combinations influenced predictive performance, we trained and tested random forest models using all possible combinations of two to eight antigens from the remaining eight antigens. Model performance was compared using AUC values (**Figure 2B**) and ROC curves (**Figure 2C**). This approach allowed assessment of both the individual discriminatory power of each antigen and the additive value of combining multiple antigens. For each panel size (two to eight antigens), different marker combinations produced similar AUCs (**Table S2**), indicating that many sets of markers performed similarly, with PvRBP2b consistently chosen in each top combination and showing the highest relative importance across models (**Table 3, Figure S1A**). There was an increase in AUC from two to five antigens, with diminishing returns in AUC as more markers were added to the model (**Figure 2B, Table 3**). The ROC curves for each top combination of *P. vivax* serological exposure markers demonstrated that the relationship between sensitivity and specificity were similar and progressively improved as the number of markers increased (**Figure 2C**). The final AUC with eight *P. vivax* antigens is 0.875 (SE: 0.004), indicative of good classification performance (**Table 3**).

**Table 3.** Top combinations from the 2 to 8 *P. vivax* serological exposure markers random forest classification models.

| SEMs | Top Combination | AUC | SE |
| --- | --- | --- | --- |
| 2 | PvRBP2b + PvMSP1-19 | 0.841 | 0.005 |
| 3 | PvCSS + PvRBP2b + PvMSP1-19 | 0.862 | 0.004 |
| 4 | PvCSS + PvRBP2b + PvMSP1-19 + PvMSP8 | 0.867 | 0.004 |
| 5 | PvPTEX150 + PvCSS + PvRBP2b + PvMSP1-19 + PvMSP8 | 0.870 | 0.004 |
| 6 | PvEBPII + PvPTEX150 + PvMSP5 + PvRBP2b + PvMSP1-19 + PvMSP8 | 0.873 | 0.004 |
| 7 | PvEBPII + PvPTEX150 + PvCSS + PvMSP5 + PvRBP2b + PvMSP1-19 + MSP8 | 0.874 | 0.004 |
| 8 | PvEBPII + PvPTEX150 + PvCSS + Pv-fam-a + PvMSP5 + PvRBP2b + PvMSP1-19 + PvMSP8 | 0.875 | 0.004 |

Abbreviations: SEM = serological exposure markers; AUC = Area under the Receiver Operator Characteristic Curve, SE = standard error.

### 3.3 Final Eight-Antigen *P. vivax* Model Demonstrates High Predictive Performance

Using the final eight *P. vivax* serological exposure markers, we tuned the random forest using Bayesian optimisation. This identified optimal hyperparameters of two for the number of predictors randomly sampled at each split (mtry) (Figure S1B) and seven for the minimum node size (Figure S1C). The final random forest model was trained with 10-fold cross-validation and five repeats to determine the optimal classification threshold and corresponding sensitivity and specificity values. The balanced random votes threshold achieved 81% sensitivity and 81% specificity (Figure 2C). Averaging across the cross-validation results, 780 individuals were classified as recently *P. vivax* infected (“seropositive”) with 366 also *P. vivax* PCR positive during the previous nine months, while 1,769 of 1,855 “seronegative” individuals had no detected *P. vivax* infection by PCR during the previous nine months. The positive predictive value (PPV) was 0.469 and the negative predictive value (NPV) was 0.954 (**Figure 2D**).

To investigate how the random forest model performed across infection categories, sex, age, and cohorts, we fitted a binomial general additive model with correct *P. vivax* serostatus classification based on *P. vivax* qPCR positive results within the previous nine months as the outcome. There was no evidence of an association between classification performance and sex (odds ratio (OR)=1.21 [95% confidence interval (CI): 0.98-1.5], *p*=0.079), but there was with age (adjusted degrees of freedom=5.16, *X*^*2*^=146.4, *p*<0.001) (Table S3). Consistent with greater sampling of younger individuals across cohorts (due to the Solomon Islands being exclusively children <12 years old) (Figure S2A), the predicted probability of correct classification was higher in younger (<14 years) than older individuals (Figure S2B).

Classification performance also varied strongly by infection status. All negative controls were correctly classified as *P. vivax* seronegative (**Figure 3A**). Performance was lowest in individuals with older *P. vivax* infections (9-12 months ago), with predicted probabilities of a correct outcome ranging from 35% in the Solomon Islands to 58% in Thailand (**Figure 3B**). By comparison, individuals with no *P. vivax* infection in the study, and those with recent (<9 months) or current *P. vivax* infections, had substantially higher probabilities of correct classification (62-81%, 68-84%, and 82-92% respectively) (**Figure 3B**). This pattern is mirrored in the odds ratios, where the odds of a correct response was markedly reduced in the old *P. vivax* infection group (OR=0.12 [95% CI: 0.048-0.30]) relative to current *P. vivax* infections, while the reduction was smaller in the recent (OR=0.46 [95% CI: 0.215-0.99]) and never *P. vivax* infected group (OR=0.367 [95% CI: 0.181-0.745]) (**Table S3**). These results show that performance is robust shortly after or without *P. vivax* infection but wanes 9-12 months post-infection.

**Figure 3.**
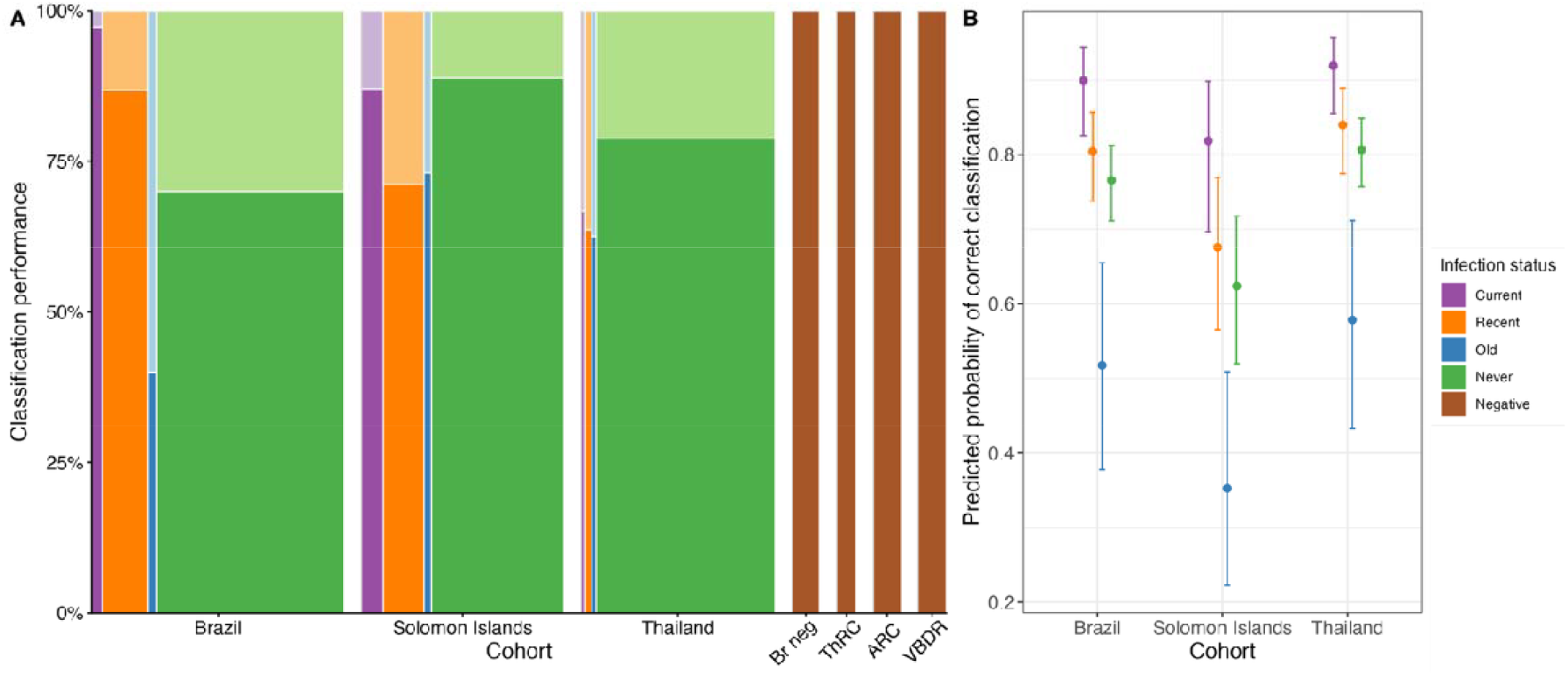
Breakdown of the classification performance of the random forest algorithm with balanced sensitivity and specificity of 81% compared to *P. vivax* qPCR reported prevalence for each cohort. **(A)** The size of each rectangle is proportional to the number of samples in each category, the coloured area represents the proportion correctly classified, while the shaded area represents the proportion mis-classified. (B) The adjusted probability of a correct classification based on a binomial general additive model (GAM) across *P. vivax* infection categories, sex, age, and the Brazil, Solomon Islands and Thailand cohorts. For the negative control cohorts, as there was no age provided for the Thai Red Cross (ThRC) or Brazil Donor Registry (Br neg) and all were correctly classified as *P. vivax* seronegative in the random forest, they were not incorporated into the model.

### 3.4 Tree-Based Classifiers Perform Marginally Better Than Other Machine Learning Models And Contributes The Most To An Ensemble Method

Random forest performance was compared with seven alternative machine learning classifiers using concurrent hyperparameter tuning and 10-fold cross validation with five repeats (**Table S4**). Decision-tree based methods (random forest, XGBoost) achieved the highest AUCs, with the random forest remaining the top classifier. Overall, AUC values differed by less than 0.07 across models, indicating only minor variation in predictive performance between algorithms (**Figure 4A**). ROC curves showed overlapping sensitivity and specificity among models, except for Naïve Bayes, which performed lowest (**Figure 4B**). An ensemble model combining all eight algorithms was then constructed to test whether it outperformed the random forest. The ensemble retained the top four classifiers (**Figure S3A**), with the random forest contributing the highest stacking coefficient (weight = 3.96) compared to XGBoost (weight = 0.95), Quadratic Discriminant Analysis (weight = 0.60) and k-Nearest Neighbours (weight = 0.59) (**Figure S3B**). The ensemble gave an AUC = 0.874 (SE: 0.0037), below the random forest and XGBoost algorithms (**Figure S3C**).

**Figure 4.**
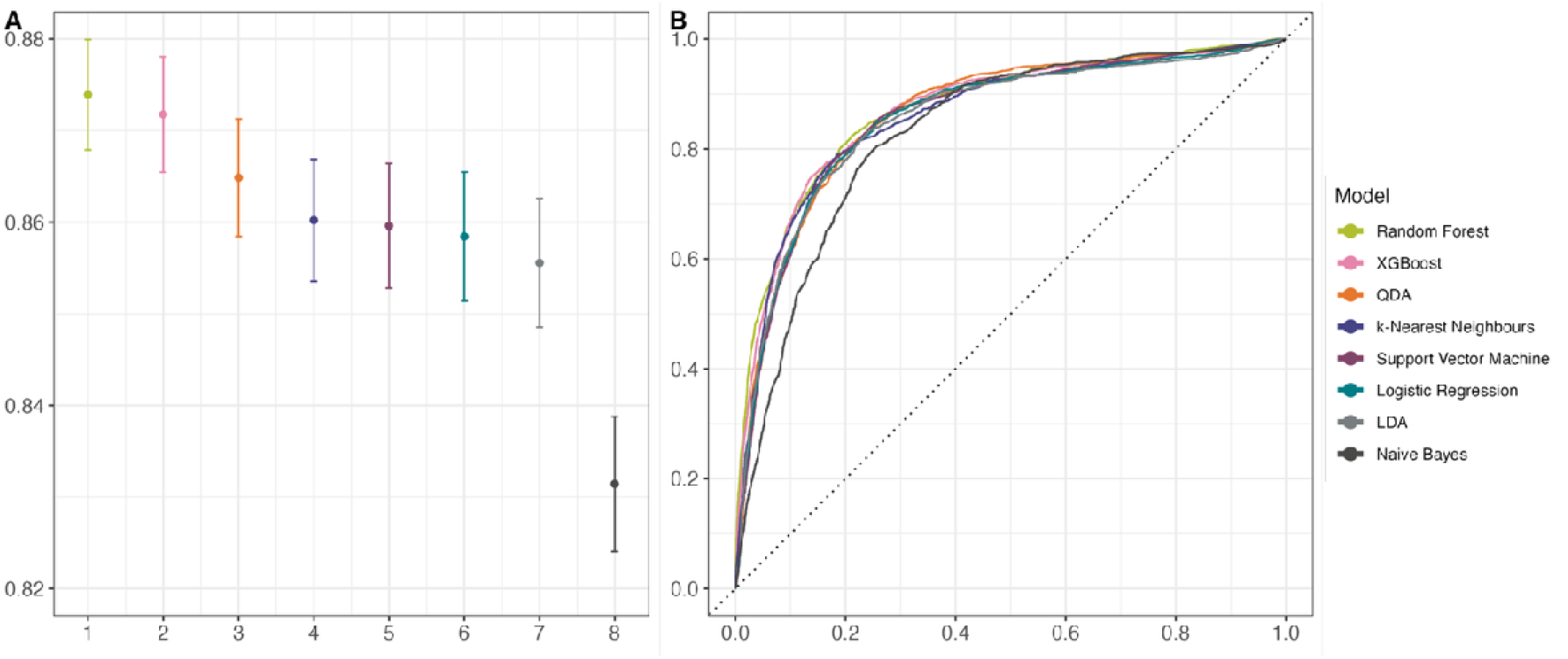
Comparison of eight supervised machine learning classification models for *P. vivax* serostatus. **(A)** AUC values for each model, ranked by the workflowsets R package. (B) ROC curves for all models, illustrating the differences in sensitivity and specificity across classification approaches.

The eight machine learning classification models and the ensemble model were evaluated using an independent validation dataset from Indonesia^41^ . The tree-based and stacked ensemble models had the strongest performance, with XGBoost (AUC = 0.937), the stack ensemble (AUC = 0.932) and random forest (AUC = 0.929) achieving the highest AUC (**Figure S4A**). Across all models, AUC values varied by 0.046, with largely overlapping ROC curves further demonstrating the marginal difference in predictive performance between algorithms (**Figure S4B**). The consistently strong performance of the tree-based models supports their use as the preferred classifiers.

### 3.5 Removing Potential Cross-Reactive Markers With Other *Plasmodium* Species Influences Final Model Performance Metrics

To account for potential cross-reactivity with other *Plasmodium* species co-endemic in *P. vivax* low transmission settings, we developed modified *P. vivax* algorithms excluding markers known to cross-react with *P. knowlesi*^*42*^ (Bourke et al. *in preparation*), *P. falciparum* or *P. malariae* (**Figure S5, Table S5**). Cross-reactive proteins with *P. vivax* were defined using two thresholds, ≥5-fold (conservative, **Figure 5**) and ≥10-fold (less conservative, **Figure S6**) above the day 7 seropositivity cut-off (**Table S6**), calculated as the mean of the malaria-naïve negative control (n=359) RAU values plus two standard deviations. Each model was trained using 10,000 trees, mtry and minimum nodes of 2 and 7 respectively, and 10-fold cross-validation with five repeats.

**Figure 5.**
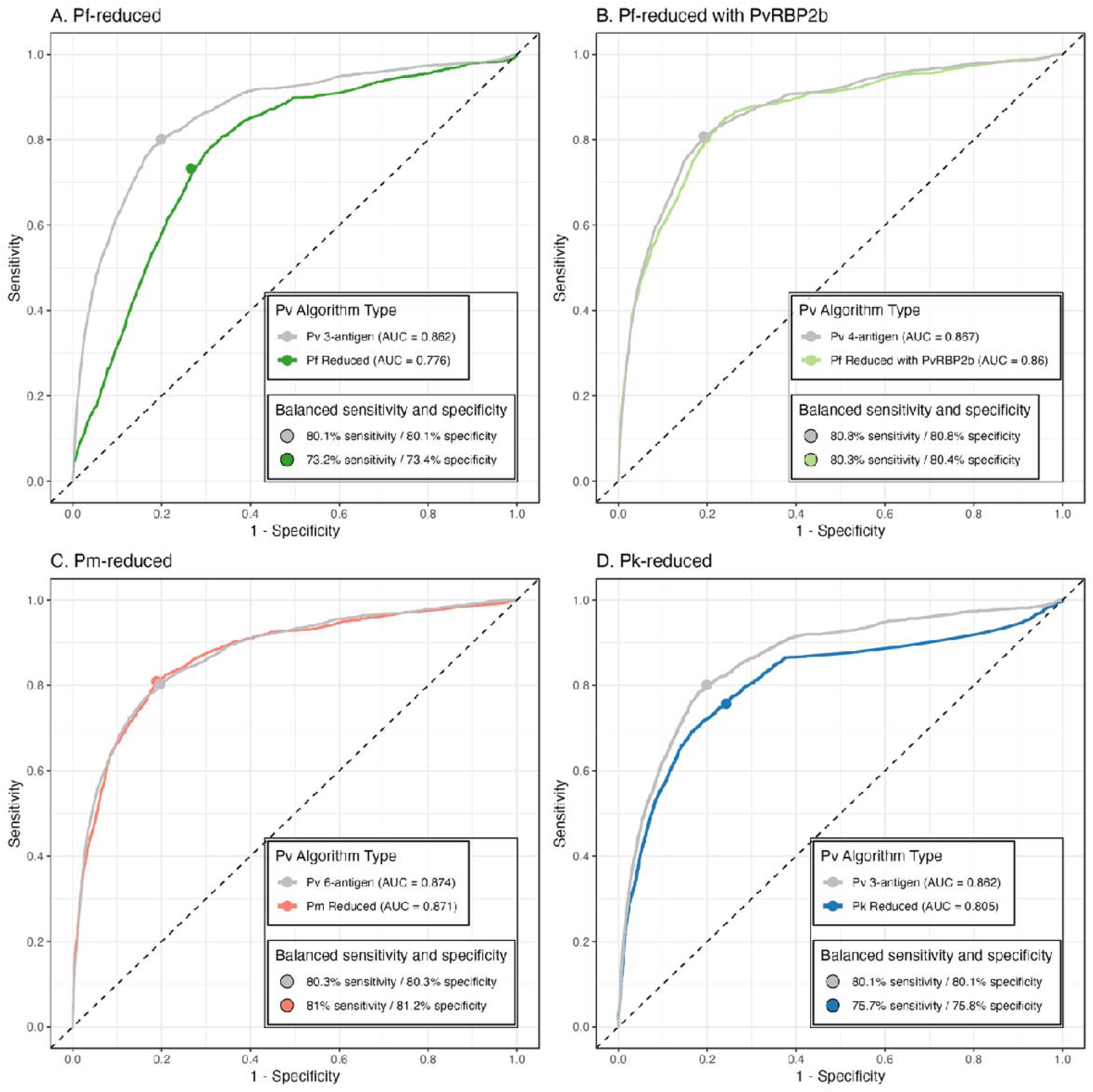
Comparison of the top performing *P. vivax* serological exposure classifiers minimising cross-reactivity with co-endemic *Plasmodium* species. ROC curves are shown for the top-performing PvSEM antigen combination (grey) alongside antigen combinations defined to minimise reactivity with markers associated with *P. falciparum* (Pf, greens), *P. knowlesi* (Pk, blue) and *P. malariae* (Pm, pink). A conservative threshold of ≥ 5-fold-change above the seropositivity cut-off was used to define antigens cross-reactive with *P. vivax*. (A) The Pf-reduced classifier includes PvMSP1-19, Pv-fam-a and PvMSP8 (dark green), while the (B) Pf-reduced classifier with PvRBP2b additionally incorporates PvRBP2b (light green). PvRBP2b is the most important antigen in the classifier and had 22.36-fold cross-reactivity. (C) The Pm-reduced classifier includes PvMSP1-19, Pv-fam-a, PvMSP5, PvCSS, PvEBPII and PvRBP2b (pink), and (D) the Pk-reduced classifier includes PvMSP5, PvEBPII and PvRBP2b (blue). The points on the ROC curve indicate the balanced sensitivity and specificity. The Area under the ROC curve (AUC) values are shown for each classifier. Each random forest classifier is run with 1,000 trees and default hyperparameters.

Using the most conservative definition of the seropositivity cut-off, the *P. falciparum*-reduced model achieved a sensitivity and specificity of 73.2%/73.4% (80.3%/80.4% when including PvRBP2b), while the *P. malariae*- and *P. knowlesi*-reduced models achieved a sensitivity and specificity of 81%/81.2% and 75.7%/75.8% respectively (**Figure 5**). When compared with the top-performing *P. vivax* antigen panels of equivalent size, classification performance (AUC, sensitivity and specificity) was largely preserved for the *P. falciparum*-reduced model with PvRBP2b and *P. malariae*-reduced model. However, there was a large drop in predictive performance in the *P. falciparum*-reduced model, driven by the removal of PvRBP2b, the strongest single-antigen classifier (**Table S1**) and most influential feature in the random forest (**Figure S1**). The *P. knowlesi*-reduced model also had a decrease in predictive performance (**Figure 5**), illustrating that a different combination of three antigens in smaller panel sizes results in lower performance compared to the top performer (**Figure S1**). All models performed marginally better when using the less conservative definition of seropositivity likely due to the inclusion of more antigens (**Figure S6**). Collectively, these findings highlight a critical trade-off whereby the *P. malariae*-reduced model remains largely comparable to the equivalent-sized top-performing panel, whilst *P. falciparum*- and *P. knowlesi*-reduced models incur substantial compromises to predictive performance.

### 3.6 PvSeroApp: A User-Friendly R Shiny Application To Process Raw Data And Apply The Classification Algorithm

The PvSeroApp was designed to streamline the data processing stages and provide an easy-to-use pipeline to implement the classification algorithm without any coding experience (Argyropoulos et al. *in preparation*). We successfully developed an R Shiny App with a GUI shown in **Figure 6A**, where users can input several key components: the experiment name, date, experiment notes, the Luminex platform used (Bio-Plex, MAGPIX® or INTELLIFLEX®), raw input files and plate layout files. Once these inputs are in the application, the user can direct themselves to the quality control tab to view plots of the standard curve, bead counts per well, MFI of the blank wells, and perform the 5-parameter logistic regression to convert MFI to RAU and visualise the plots (which should be linear). Once the user is satisfied with the quality of their data, they can progress to the final tab to choose the classification algorithm, select the balanced sensitivity and specificity threshold of 81% each or a 90% specificity threshold (with 67.5% sensitivity) based on the training dataset, and run the classification algorithm on their data. The page immediately shows a table of counts for seropositive and seronegative samples per plate in the dataset. At each stage, the user can save the datasets used for these analyses and summary statistics. The PvSeroApp has been successfully tested with collaborators across 20 countries as of May 2026 (**Figure 6B**).

**Figure 6.**
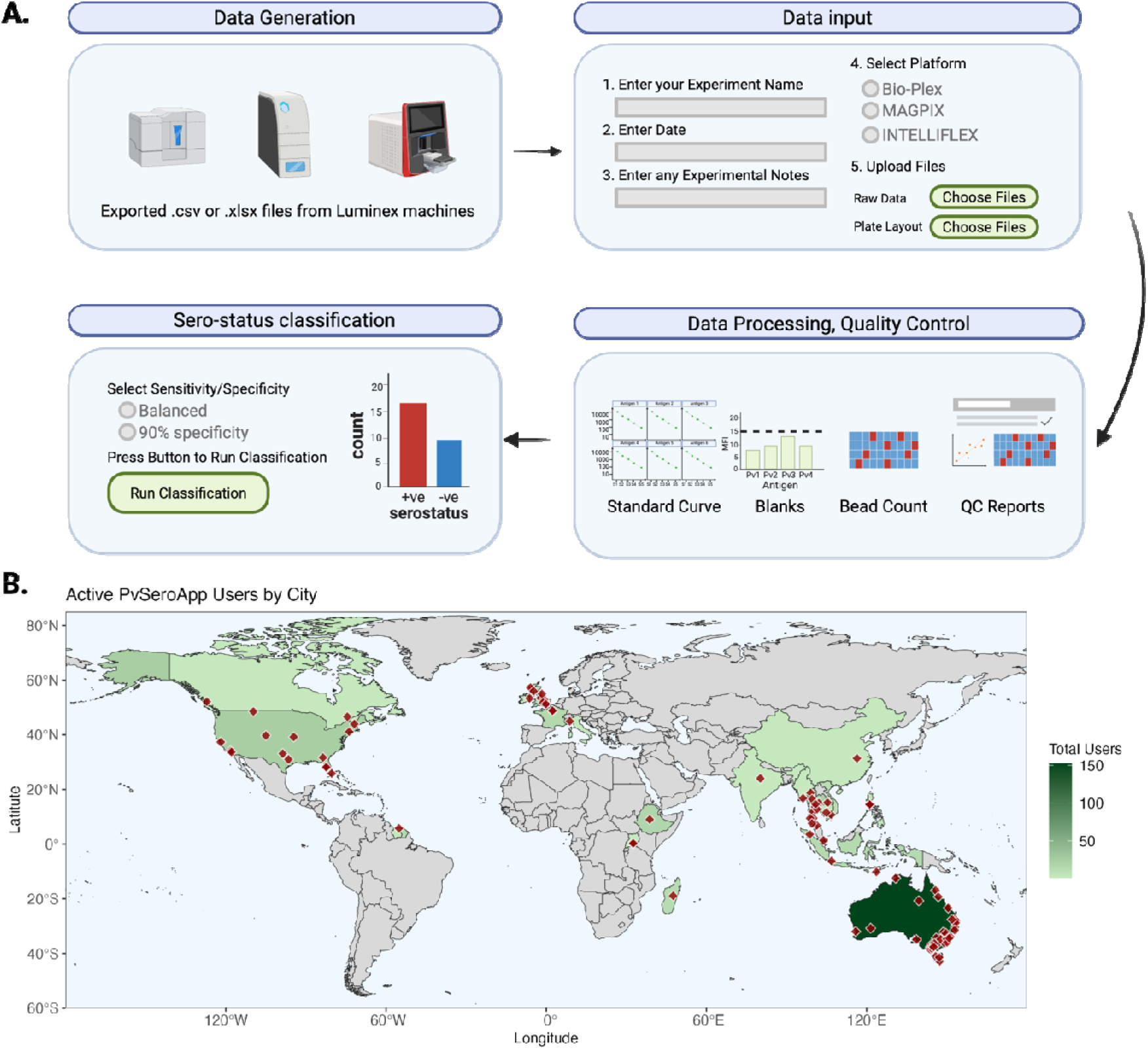
The PvSeroApp: A fit-for-purpose tool to support *P. vivax* sero-surveillance via serological data processing and statistical analysis. **(A)** Overview of the workflow from raw Luminex data input, through processing and quality control, to final classification using a random forest algorithm. (**B**) Geographic distribution of PvSeroApp users as of May 2026 with active cities (red) and countries (green) highlighted. Figure created with BioRender.com.

## 4 Discussion

Here we present a random forest classification algorithm using eight serological exposure markers to classify serostatus, as a measure of recent exposure to *P. vivax*. Markers were chosen that balance the assay complexity and scalability with optimal subsets that maintained algorithm performance. This work presents the first major update to the assay and algorithm since 2020. The top-performing combination of eight antigens for classifying serostatus achieved an AUC of 0.874 at a balanced voting threshold of 81% sensitivity and 81% specificity. The final random forest algorithm consistently outperformed alternative machine learning classification algorithms, with marginal differences in performance metrics, and contributed the greatest weight to a meta-ensemble method, rendering the random forest the most appropriate classification model for these data. Removing potential cross-reactive antigens ultimately resulted in similar, albeit slightly diminished, classification performance. An online R Shiny application (PvSeroApp) was developed to automate raw serological data processing, quality control and serostatus classification, and facilitated PvSeroTAT implementation in multiple malaria-endemic countries. Ultimately, the PvSEM assay and algorithm combination offers a sero-surveillance strategy to target anti-hypnozoite therapy and strengthens the toolkit for *P. vivax* elimination.

The updated multi-antigen panel used in the machine learning classifier for identifying recent *P. vivax* infections demonstrates performance comparable to our 2020 model^12^, suggesting that the approach may be nearing a performance plateau imposed by the reference standard rather than limitations in antigen selection or model design. However, our current panel includes antigens with single-antigen AUC values ranging from 0.729 (PvCSS) to 0.832 (PvRBP2b_169-470_), representing an improvement over the previous iteration, in which AUCs ranged from 0.670 (PvMSP3.10) to 0.816 (PvRBP2b_161-1454_ )^12^. These improvements likely reflect advances in both antigen^20,43^ and assay^44^ design. The revised panel excludes PvMSP3, which was identified as large and unstable^19,20^, and PVX_112670 (unknown function) and PvRAMA due to poor performance. It instead incorporates PvCSS and PvMSP5, both highly immunogenic antigens implicated in merozoite invasion of reticulocytes^33,39,42,45^, and PvPTEX150 of unknown function in *P. vivax* although its ortholog in *P. falciparum* is expressed in late schizogony and is essential to blood-stage development^38,42^. In addition, for each panel size (two to seven antigens), different marker combinations resulted in a redundancy in classification performance, and PvRBP2b was essential as it was consistently chosen in the top combination in each panel size, as also shown in Sa et al.^46^. This robustness is advantageous for practical deployment, as it allows antigen selection to be tailored to specific use-cases and production constraints, conferring a high degree of modularity and flexibility.

We evaluated eight machine learning algorithms and one meta-learner, with tree-based methods consistently performing the best. Overall, the models had AUCs spanning a narrow range and near-overlapping ROC curves, indicating substantial redundancy among modelling approaches for these data. Although the random forest achieved the highest performance on the training dataset, XGBoost marginally outperformed other models on the validation dataset, with the stack ensemble and random forest within 0.008 AUC. We ultimately selected the random forest due to its greater robustness to overfitting^47,48^. Unlike XGBoost, which builds sequential, interdependent trees^49^, random forests rely on bootstrap aggregation and feature randomness to stabilise predictions through averaging across independent trees, making random forests less prone to overfitting^48,50,51^. Given that the model was trained on data from only three P. vivax-endemic settings, prioritising robustness and generalisability over marginal performance gains was considered essential. However, XGBoost’s computational efficiency^48,49^ makes it more suitable for other PvSEM applications operating under strict memory or processing constraints, such as rapid diagnostic tests.

As *P. vivax* rarely circulates in isolation in a population, we developed species-specific reduced models to evaluate the impact of cross-reactivity on diagnostic performance. *P. malariae* infections are often chronic and co-occur with other species^52,53^, and because they are not always treated promptly, they may influence assay performance despite lower genetic similarity to *P. vivax*^*54*^. The *P. malariae*-reduced model maintained similar performance to the top performing equivalent *P. vivax* panel (AUC = 0.871), demonstrating that cross-reactivity can be addressed without sacrificing performance. In contrast, when we account for potential cross-reactivity with *P. falciparum*, the major malaria species that frequently co-occurs with *P. vivax*, the model exhibits substantial performance loss (AUC=0.776, sensitivity=72.3%, specificity=73.4%), relative to the top performing classifier of equivalent size, primarily due to the absence of PvRBP2b, the strongest contributor to the random forest classifier. Retaining PvRBP2b in the *P. falciparum*-reduced model improved performance and was comparable to the *P. vivax* panel of a similar size (AUC=0.867, sensitivity=80.3%, specificity=80.4%). *P. knowlesi*, which shares the highest genetic similarity with P. vivax, presented a more complex challenge. Our conservative cross-reactivity threshold yielded a three-antigen *P. knowlesi*-reduced model with substantially lower performance (AUC=0.805, sensitivity=75.7%, specificity=75.8%), than the top *P. vivax* combination of equivalent panel size. Notably, this three-antigen panel performs worse than the eight-antigen *P. knowlesi*-reduced model from Longley et al.^40^ (AUC=0.840, sensitivity=78.7%, specificity=76.9%), which may reflect differences in panel size or cross-reactivity thresholds. Longley et al.^40^ previously classified PvPTEX150 as non-cross-reactive, whereas our conservative threshold now defines it as cross-reactive. Applying a less-conservative threshold to include PvPTEX150 did not substantially improve performance relative to the equivalent top performing *P. vivax* panel. Critically, no antigens were entirely non-cross-reactive with at least one species. Given the poorer performance of *P. falciparum*- and *P. knowlesi*-reduced models compared to *P. vivax* panels of equivalent size, we recommend using the full eight-antigen *P. vivax* panel with species-specific PCR confirmation rather than relying on reduced models with compromised accuracy.

In practice, the PvSEM machine learning classification algorithm can be tailored to three operational scenarios. The first uses a balanced threshold (81% each) which optimises both sensitivity and specificity. The second prioritises a high sensitivity with lower specificity (95% sensitivity, 42.4% specificity) to maximise the identification and treatment of all likely hypnozoite carriers at the cost of some over-treatment. The third prioritises high specificity with lower sensitivity (52.4% sensitivity, 95% specificity) suited for surveillance or conservative settings where over-treatment is not tolerated. In this scenario, the aim is to correctly identify non-carriers and minimise treatment of uninfected individuals, while the impact of false negatives can be mitigated by increasing survey size^12^. As radical cure carries a risk of haemolysis in G6PD-deficient individuals^10^ and G6PD screening can add a substantial burden to healthcare costs^55–57^, operational use of a high-sensitivity, low-specificity approach is unlikely. A modelling study of PvSeroTAT deployment in Brazil indicates that adjusting the sensitivity and specificity following one round can substantially influence population-level outcomes, with increases in either predicted to reduce *P. vivax* PCR prevalence by approximately 25% depending on the location^17^. To facilitate real-world application, we developed an R shiny interface that allows users to apply the random forest model without coding expertise, supporting end-to-end use of the PvSeroTAT pipeline and sensitivity and specificity threshold strategies according to local epidemiological priorities, such as choosing higher specificity in near-elimination settings.

Overall, this work strengthens the versatility of the PvSEM assay and algorithm, offering a modular tool adaptable to different epidemiological contexts and deployment strategies. By reliably detecting recent *P. vivax* exposure and identifying individuals at risk of carrying hypnozoites, this assay and algorithm enable more targeted and efficient interventions, potentially reducing overtreatment and resource use compared with blanket strategies such as mass drug administration. Combined with the R Shiny interface, this updated assay and algorithm provide a robust tool for mapping recent *P. vivax* exposure and guiding targeted public health interventions.

### Limitations of the study

The PvSEM machine learning algorithm was designed, developed and trained using datasets from low-transmission *P. vivax* settings. Consequently, the current eight-antigen panel has not yet been validated across a range of epidemiological contexts, such as moderate- and high-transmission settings. This limitation is reflected in the model’s predictive performance: in low transmission settings, where prevalence is typically below 10%^58^, a lower PPV is expected due to the increased proportion of false positives that naturally arise when infection rates are low. Conversely, the high NPV demonstrates strong reliability in identifying uninfected individuals, supporting the use of PvSeroTAT strategy and/or for surveillance in elimination-focused applications.

Our analysis of cross-reactivity was focussed on *P. falciparum* or *P. malariae* clinical samples from Sabah, Malaysia, at a time when the region had low levels of *P. vivax* transmission. Cross-reactivity patterns in settings with higher *P. falciparum* endemicity and emerging *P. vivax* transmission, such as Ethiopia and Madagascar, may therefore differ from those observed here and may be more complex to disentangle due to the similar risk-factors for both species. This requires further evaluation but may also have limited programmatic importance depending on the specific use-case and epidemiological setting.

We also identified age as a factor influencing classification performance. This effect can largely be attributed to over-sampling younger children in our training datasets. Immune and parasite exposure profiles differ with age and are interrelated^59^. Future work will further investigate age-dependent variation and its implications for model performance and generalisability. Despite age being identified as an influencing factor, the algorithm performed well with an AUC of 0.92 in a validation dataset of Indonesian soldiers aged between 18 and 65^41^.

Finally, while this study used an eight-antigen panel, our analyses demonstrate that similar classification performance can be achieved using a reduced set of three to five antigens. Ongoing research aims to quantify how reducing the number of antigens to three to five may impact the PvSeroTaT approach in different transmission contexts.

## Data Availability

All data produced are available online at https://github.com/dionnecargy/SeroTrackR.

## 5 Materials and Methods

### 5.1 Key Resources Table

| REAGENT or RESOURCE | SOURCE | IDENTIFIER |
| --- | --- | --- |
| Antibodies |  |  |
| PE-conjugated Donkey F(ab)2 anti-human IgG | Jackson ImmunoResearch | JIR 709-116-098 RRID:AB_2340519 |
| Chemicals, peptides, and recombinant proteins |  |  |
| 4 *P. vivax* proteins expressed in *E. coli* | Zip Diagnostics |  |
| 1 *P. vivax* peptide | Genscript |  |
| 1 *P. vivax* protein expressed in *E. coli* | Tham Laboratory, WEHI |  |
| 1 *P. vivax* proteins expressed in mammalian cells | WEHI Protein Production Facility |  |
| 3 *P. vivax* proteins expressed in insect cells | WEHI Protein Production Facility |  |
| Software and algorithms |  |  |
| PvSeroApp R Shiny App | Argyropoulos et al. *in prep* | https://github.com/dionnecargy/PvSeroApp |
| SeroTrackR | Argyropoulos et al. *in prep* | https://github.com/dionnecargy/SeroTrackR |
| Other |  |  |
| MAGPIX Instrument | Luminex | https://www.luminexcorp.com/magpix-system/#overview |

### 5.2 Resource Availability

#### 5.2.1 Lead contact

Further information and requests for resources should be directed to and will be fulfilled by the lead contact, Dr Rhea Longley.

#### 5.2.2 Materials availability

This study did not generate new unique reagents. Access to the Luminex assay reagents can be requested through the Vivax Serology Partnership (vispa.online).

#### 5.2.3 Data and code availability

All data are available in the main text or GitHub repositories, along with the code, at: https://github.com/dionnecargy/PvSeroApp or https://github.com/dionnecargy/SeroTrackR. Any additional information required to reanalyse the data reported in this paper is available from the lead contact upon request.

### 5.3 Method details

#### 5.3.1 Study design and Molecular Assay Data

To assess antigen-specific IgG antibody responses towards our panel of antigens, we measured these responses in plasma samples from people resident in malaria-endemic regions of Thailand, Brazil and Solomon Islands. Importantly, new antigen constructs and expression batches as well as a modified Luminex assay were utilised compared to the data published in Longley et al^12^. Briefly, year-long observational cohort studies were conducted in Kanchanaburi and Ratchaburi provinces in Thailand^60^ (neighbouring provinces with similar epidemiological profiles, treated as one population), Manaus, Brazil^61^, and Ngella, Solomon Islands between 2013 and 2014^62^. We also included *P. vivax* negative controls from the Australian Red Cross (ARC), Brazil Blood Donor Registry (Br Neg), Thai Red Cross (TRC), and the volunteer biospecimen donor registry (VBDR) in Victoria, Australia. **Table 2** outlines the number of samples included from malaria-endemic regions and negative controls.

Individuals enrolled in the year-long cohort studies provided a blood sample every month, which was screened by light microscopy and qPCR for the detection of blood-stage *P. vivax* infection, allowing us to determine which individuals were infected with *P. vivax* and at which time point during the year-long studies. At any time point, individuals with a fever (body temperature ≥37.5°C) and tested positive for malaria from a rapid diagnostic test (SD BIOLINE Malaria Ag P.f./Pan, Standard Diagnostics, Republic of Korea (Thailand), CareStart, Access Bio, USA (Solomon Islands)) or thick blood smear (Brazil), they were referred to their local malaria clinic for treatment as per national guidelines (this was not monitored by the study team). Individuals were defined as currently infected if they tested positive for *P. vivax* by light microscopy or qPCR in the final visit (visit 12) of the 12-month cohort survey, and recently infected between months 3 to 12 of the survey. These individuals were not treated as the study was observational. Those who tested positive only in months 1 or 2, or who had no positive result during the survey period, were classified as not recently infected with P. vivax. This definition reflects that an individual will likely experience a relapse within nine months following a primary blood-stage infection with tropical and sub-tropical *P. vivax* strains.

Antigen-specific total IgG antibody responses were measured using a multiplexed Luminex® bead-based assay (Bareng et al. *in preparation*) at the final visit of the year-long study. This allowed us to characterise antibody responses in relation to time since prior *P. vivax* infection. All plasma samples from the 2,635 isolates were assayed at a 1:100 dilution. Median fluorescent intensity (MFI) values obtained from the Luminex® MagPix were converted to Relative Antibody Units (RAU) using a standard curve generated with pooled positive control plasma from highly immune adults from Papua New Guinea (PNG)^63^. The standard curve was run on every 96-well plate. A five-parameter logistic function was applied to calculate equivalent dilution values relative to the PNG control plasma (ranging from 1.95 x10^-5^ to 0.02), and the conversion was implemented as the “MFItoRAU” function in the SeroTrackR R Package (https://github.com/dionnecargy/SeroTrackR) (Argyropoulos et al., *in preparation*). Other positive control plasma pools can be used, provided they are run side by side with the PNG pool (see supplementary information for example; standard curve concentrations from 1.95 x10^-5^ to 0.02). As the random forest algorithm, described below, is trained on PNG-derived RAU values, a custom MFI-to-RAU conversion function must be generated by directly comparing the alternative pool to the PNG pool, ensuring that the RAU is equivalent before applying the model.

#### 5.3.2 Selection of Serological Exposure Markers

Previous work characterised antibody responses against 307 *P. vivax* proteins over time and identified several candidate serological markers for recent *P. vivax* exposure^64,65^. Here, we extend these findings through iterative improvements of the antigens, assay and algorithm. Fourteen *P. vivax* serological exposure markers were initially selected by combining the top eight markers from the first PvSeroTAT algorithm^12^ with additional high performing candidates identified later^42^ (**Figure 1A**). In doing so, we balance the selection of serological exposure markers that are associated with high classification performance, with the selection of proteins that are easier to manufacture and are more stable. Several candidates were subsequently excluded: PvMSP3a due to its large size (828 amino acids) and genetic complexity^19,20^; Pv-fam-a (PVX_112670) and PvRipr due to expression difficulties; PvRBP2a due to high genetic diversity^20^; and Pvs16 and PvRAMA due to low classification performance (AUC) (**Figure 1A**).

#### 5.3.3 Classification algorithms

Individuals were classified as “sero-positive” (recently *P. vivax* infected) or “sero-negative” (not recently *P. vivax* infected) depending on their *P. vivax* blood-stage infection history in the past nine months. Infection status, determined by qPCR, served as the binary outcome variable, while RAU values for each serological exposure marker were used as predictor variables in the machine learning algorithms described below. To maximise simplicity and broad applicability, we restricted the model inputs to antibody measurements and did not include age or sex, minimising the dependence on supplementary data and enhancing practical usability.

##### 5.3.3.1 Single antigen classifications

We calculated the single-antigen classification performance for each antigen considered in our panel, calculating sensitivity and specificity thresholds, and generating ROC curves to evaluate the predictive performance of each antigen in distinguishing recent and not recent *P. vivax* infections (i.e., infection within the prior nine months). We created 1,000 cut-off thresholds between zero and the maximum RAU for each antigen, and calculated the corresponding sensitivity 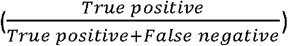 and specificity 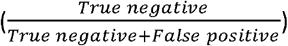 for each cut-off threshold.

##### 5.3.3.2 Random forest classification models

To explore the top combinations, we trained a random forest on all possible combinations of the eight final eight *P. vivax* serological exposure markers as described in **Figure 1A**. We created a random forest classification algorithm for all possible combinations of eight *P. vivax* markers, using 10-fold cross validation with five repeats. For this, we fit the random forest using the ranger v0.17.0 R package^66^ in R v4.4.1^67^, with 1,000 trees, and all other hyperparameters as default. The top-performing combination of two to eight *P. vivax* markers were evaluated using the AUC.

Using our top combination of eight *P. vivax* antigens, we first evaluated the importance of each variable that contributes to the random forest, calculated using the vip R package v0.4.5^68^. We performed Bayesian optimisation to tune the hyperparameters mtry, the number of predictors randomly sampled at each split, and minimum node size (min_n), the minimum number of observations required in a terminal node. We specified 1,000 trees, and permutation-based variable importance for optimisation of mtry and minimum node size. Search ranges were defined for the number of predictors at each split (mtry: 1 to total number of predictors) and the minimum node size (min_n: 1 to 40). To initialise the Bayesian optimisation, we used a 50-point Latin hypercube design, evaluated using 10-fold cross validation and AUC. Bayesian optimisation then proceeded for 300 iterations, with early stopping after 100 iterations without improvement. The optimal hyperparameters were selected based on maximum cross-validated AUC.

The final model is trained on the whole dataset, with 10,000 trees and the optimised mtry and minimum node size. The final AUC is calculated, as well as sensitivity, specificity, the positive predictive value 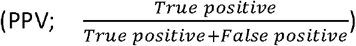 and negative predictive value 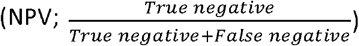.

A binomial generalized additive model with a logit link was fitted to estimate adjusted odds ratios for a correct classification response. The model included *P. vivax* infection status, self-reported sex, cohort, and a penalised cubic regression spline for self-reported age, implemented using the “gam” function in the mgcv R package (v1.9-3; R Core Team, Austria). A correct response was defined as either (i) *P. vivax* seropositive with qPCR-confirmed *P. vivax* positive result within the previous nine months, or (ii) *P. vivax* seronegative with no qPCR-detected *P. vivax* infection within the previous nine months. Adjusted odds ratios were estimated using the emmeans R package (v2.0.0)^69^. Adjusted predicted probabilities by cohort and infection status were visualised using the ggeffects R package (v2.3.1)^70^. Individuals with missing data were excluded from the analyses, including all individuals from the Brazil Blood Donor Registry and Thai Red Cross, as age and sex were unknown (n = 189; 7% of dataset). The proportion of correct *P. vivax* serostatus classification by cohort and infection status was visualised using ggplot2 (v4.0.1)^71^.

#### 5.3.4 Comparison of random forest against other machine learning models

Machine learning model calibration was performed using the workflowsets package v1.1.0^72^ within the tidymodels framework^73^. Models were implemented via the parsnip interface using the appropriate computational engines: logistic regression (stats package v4.4.0)^74^, gradient boosted trees (xgboost v1.7.11.1)^75^, k-nearest neighbours (kknn v1.4.1)^76^, linear discriminant analysis and quadratic discriminant analysis (MASS v7.3-65)^77^, naïve Bayes (naivebayes v.1.0.0)^78^, random forest (ranger v0.17.0)^66^, and support vector machines (kernlab v0.9-33)^79,80^. Hyperparameter tuning was conducted using the workflowsets object with 10-fold cross-validation and five repeats for models containing hyperparameters available to tune (gradient boosted trees, k-nearest neighbours, naïve Bayes, and support vector machines). Optimal hyperparameters were selected based on AUC performance. Final models were trained on the longitudinal cohort dataset using 10-fold cross-validation with five repeats and the optimized hyperparameters. Classification performance was assessed and ranked according to final AUC values, with additional evaluation using ROC, PPV, NPV, sensitivity, and specificity.

The stacks R package v1.1.1^81^ was used to combine the eight machine learning models into a single stacked ensemble model to assess whether a multi-algorithm ensemble improved *P. vivax* classification performance compared with individual models. All eight candidate models underwent hyperparameter-tuning using the 10-fold cross-validation with five repeats, and the final tuned versions were used as inputs to the stack ensemble. To create the ensemble, we applied a regularised linear model (LASSO) meta-learner which fits the assessment-set predictions of each candidate and assigns a stacking coefficient (LASSO beta values) that weights the contribution of each member to the final prediction. Candidates with non-zero stacking coefficients were selected as ensemble members, and these were re-fitted using the full training set. The regularisation strength used in the LASSO was selected through resampling to balance the number of contributing models and predictive performance, where higher penalties result in fewer candidates and lower penalties including more. Ensemble classification performance was first assessed by examining (i) the number of contributing models, (ii) the relative weights (stacking coefficients) of each member, and (iii) global metrics such as the Brier class and AUC. Final *P. vivax* classification performance was then evaluated on a validation dataset^41^ (see supporting information for details) using ROC analysis, AUC, sensitivity, and specificity. These same metrics were calculated for each individual machine learning model to enable direct comparison with the ensemble approach.

#### 5.3.5 Accounting for cross-reactivity with other *Plasmodium* species

We evaluated the top-performing eight-marker *P. vivax* panel against combinations excluding serological exposure markers identified as highly cross-reactive with *P. knowlesi* ^42^, *P. falciparum* or *P. malariae*. For *P. falciparum* and *P. malariae*, the data are presented here in the supplementary for the first time. Briefly, total IgG antibody responses were assayed against the *P. vivax* antigen panel in plasma collected from individuals from Sabah, Malaysia with clinical PCR-confirmed *P. falciparum* or *P. malariae* monoinfections at the time of presentation (day 0) and at 7 and 28 days after commencing antimalarial treatment, as described^82^. Sample collections occurred at a time when all four *Plasmodium* species were co-endemic in Sabah, but with low pre-elimination transmission of P. vivax, *P. falciparum* and *P. malariae*^83^. A protein-specific seropositivity cut-off was defined as the mean of the malaria-naïve negative-control (n=359) RAU values plus two standard deviations^42^ (**Table S6**). One isolate was excluded due to an outlier RAU value (>0.01) for PvCSS (**Figure 1C**). We evaluated two definitions of cross-reactivity: ≥5-fold (conservative) and ≥10-fold (less conservative) at day 7 above the seropositivity cut-off. We assessed the genetic identity and similarity between *P. vivax* Sal-1 proteins and their orthologs in *P. falciparum* 3D7, *P. malariae* UG01 and *P. knowlesi* H strains. Ortholog sequences and synteny were obtained from PlasmoDB^84^. Whole-protein alignments against *P. vivax* Sal-1 were performed using the EMBOSS Needle^85^ to calculate the percentage identity and similarity. In addition, the construct *P. vivax* sequences used in the *P. vivax* serology assay analysed using NCBI BLASTp^81^, and the top hits for 3D7 (taxid: 36369), UG01 (taxid: 5858) and H (taxid: 5851) were recorded (Table S5).

For each panel, a random forest classifier with 1,000 trees, default hyperparameters, and 10-fold cross-validation with five repeats was trained and tested. Using the more conservative definition, markers cross-reactive with *P. falciparum* were PvMSP5, PvPTEX150, PvRBP2b, PvEBP, and PvCSS (**Figure S5**; **Table S6**). Two “*P. falciparum*-reduced” algorithms were tested: one excluding these five antigens (including Pv-fam-a (PVX_096995), PvMSP1-19, PvMSP8), and another also including PvRBP2b (Pv-fam-a (PVX_096995), PvMSP1-19, PvMSP8, PvRBP2b), which is the top serological exposure marker as demonstrated previously^12^ and in this study. PvPTEX150 and PvMSP8, identified as cross-reactive with *P. malariae*, were removed to create the “*P. malariae*-reduced” panel (**Figure S5**; **Table S6**). The “*P. malariae*-reduced” classifier includes PvMSP1-19, Pv-fam-a, PvMSP5, PvCSS, PvEBPII, PvMSP8 and PvRBP2b. Markers previously identified in Longley et al.^42^ as highly cross-reactive with *P. knowlesi* were PvMSP8, PvMSP1-19, Pv-fam-a (PVX_096995), PvPTEX150 and PvCSS (**Table S6**); the corresponding “*P. knowlesi*-reduced” panel included PvMSP5, PvEBPII and PvRBP2b. It is important to note that overall, the *P. vivax* antigen panel was most highly cross-reactive in plasma from *P. knowlesi*-infected individuals and much more moderately and for a shorter duration in plasma from *P. falciparum*- and *P. malariae*-infected individuals (**Figure S5**).

#### 5.3.6 Implementing the model in an R Shiny Application

The final random forest *P. vivax* classification algorithm was encoded in the SeroTrackR R package which was initially developed to support the PvSeroApp (https://dionnecargy.shinyapps.io/PvSeroApp/) (Argyropoulos et al. in prepration). The PvSeroApp was developed using RStudio v2024.12.1+563 ^86^ leveraging the shiny v1.10.0 ^87^ and shiny.fluent v0.4.0 ^88^ packages for the user-interface and server options. The waiter v0.2.5 ^89^ was used for loading screens, spsComps v0.3.3.0 ^90^ was used for error messaging, tidyverse v2.0.0 ^91^ for data wrangling and visualisation, plotly v4.10.4 ^92^ for interactive plotting, rmarkdown v2.29 ^93,94^ for pdf output and the readxl v1.4.3 ^95^, openxlsx v4.2.7.1 ^96^, janitor v2.2.1 ^97^ for data processing and wrangling. User engagement with the PvSeroApp was monitored using Google Analytics (Google LLC, 2025), which provided anonymised data on active users by city and country. Maps were developed using the R packages ggmap v4.0.2 ^98^ and rnaturalearth 1.1.0 ^99^.

### 5.4 Additional resources

The SeroTrackR R package and updates are available via GitHub: https://github.com/dionnecargy/SeroTrackR and the PvSeroApp R Shiny Application source code is available at https://github.com/dionnecargy/PvSeroApp.

## 6 Acknowledgements

We would like to thank prior work from Dr Shazia Ruybal-Pesantez and Dr Connie Li-Wai-Suen of an automated quality control platform and standard curve conversion implemented in R. We would like to thank Dr Michael White and Dr Thomas Obadia for their feedback on the random forest model and R Shiny application based on their prior work. We also would like to thank the contributions of all study participants in the described cohort studies and negative control samples. We acknowledged proteins produced and purchased from the Tham Laboratory (Wai-Hong Tham, WEHI), the WEHI Protein Production Facility (Marija Dramicanin), and ZiP Diagnostics. We acknowledge the Cowman Laboratory (Benjamin Seager, Stephen Scally, WEHI) for initial preliminary test expressions of some proteins. We acknowledge efforts of both Natalie Senzo and Julie Healer as project managers at WEHI.

## 7 Ethics Approvals

The Ethics Committee of the Faculty of Tropical Medicine, Mahidol University, Thailand, approved the Thai year-long cohort study (MUTM2013-027-01). The Brazilian year-long cohort study was approved by the Ethics Review Board of the Fundação de Medicina Tropical Dr. Heitor Vieira Dourado (349.211/2013) and by the Brazilian National Committee of Ethics and by the Ethics Committee of the Hospital Clínic, Barcelona, Spain (2012/7306). The National Health Research and Ethics Committee of the Solomon Islands Ministry of Health and Medical Services (HRC12/022) approved the Solomon Islands year-long cohort study. The Human Research Ethics Committee at the Walter and Eliza Hall Institute of Medical Research (WEHI) approved samples for use in Melbourne (14/02) and also approved use and collection of the control panel samples (14/02). The Medical Research and Ethics Committee, Ministry of Health, Malaysia (NMRR-10-754-6684 and NMRR-19-4109-52179) and Menzies School of Health Research, Australia (HREC 2010-1431 and 2023-4617) approved the use of the Malaysian clinical samples.

## 9 Authorship

Conceptualisation – LS, DCA, IM, RL; Data Curation – LS, DCA, TW, NMA, MJG, JS, ML, VV,RM, IM, RJL; Formal Analysis – LS, DCA, APNB, KWW; Funding Acquisition - NMA, MJG, IM, RJL; Investigation - LS, DCA, APNB, JL, NKW, ML, AA, PSL, KWW, RM; Methodology – LS, DCA, APNB, JL, NKW, ML, AA, PSL, KWW, RM, IM, RJL; Supervision – IM, RJL; Visualisation – LS, DCA, APNB; Writing – Original Draft Preparation: LS, DCA, RJL; Writing – Review & Editing - LS, DCA, APNB, JL, NKW, ML, AA, PSL, KWW, TW, NMA, MJG, JS, ML, VV, RM, IM, RJL.

## 10 Supporting Information

### Positive control plasma pools

A quality assurance/quality control (QA/QC) plate containing both the PNG pool and the other positive control plasma pool of interest, for example one using magnetic beads coupled with Ethiopian plasma (ETH), were run side-by-side for the standard curve concentrations from 1.95 x10^-5^ (S10) to 0.02 (S1) ten times. Using the MFItoRAU_Adj function in the SeroTrackR R package, a five-parameter logistic function was applied to calculate the equivalent dilution values relative to the standard curves from both PNG and ETH plasma pools. As the random forest algorithm is trained on PNG-derived RAU values, this custom MFI-to-RAU conversion function (MFItoRAU_Adj) must be generated by directly comparing the alternative pool to the PNG pool, ensuring that the RAU is equivalent before applying the model (Argyropoulos et al., *in preparation*).

### Validation dataset

This study followed 592 Indonesian soldiers who had recently returned from nine-month deployments in malaria-endemic Papua to malaria-free East Java^41^. Participants provided fortnightly blood samples for six months for microscopy and PCR (in subsets), and IgG antibody measurements to the eight *P. vivax* antigens using a Luminex MAGPIX® bead-based assay. We assessed the performance of machine learning algorithms applied to antibody levels from participants at enrolment, using microscopy and PCR to determine whether soldiers went on to relapse over the six-month follow-up period.

**Figure S1.**
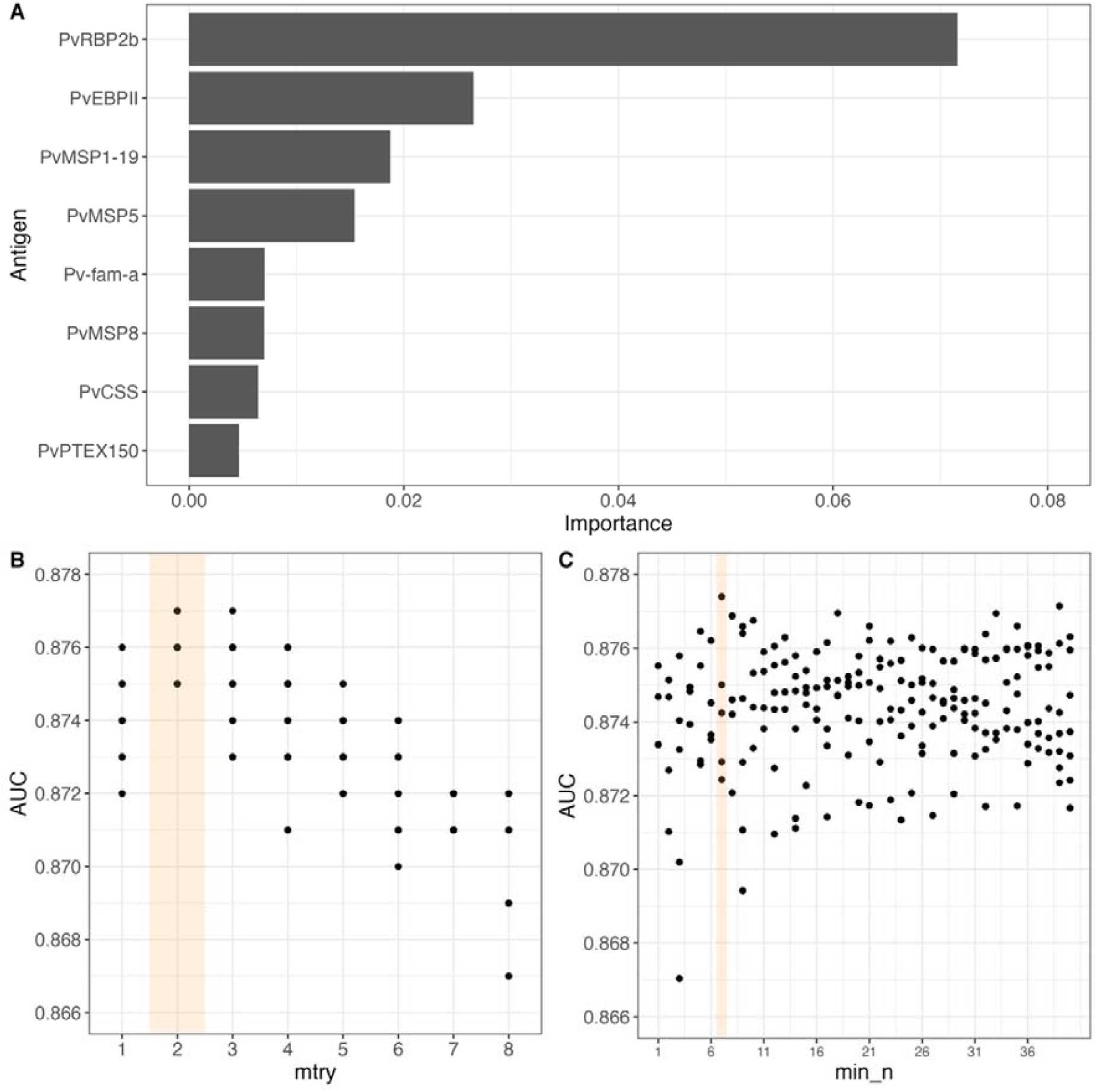
Variable importance and hyperparameter tuning outputs. (**A**) The variable importance of *P. vivax* antigens in the random forest model was calculated using the vip R package v0.4.5 ^68^. Results from hyperparameter tuning for (**B**) “mtry” and (**C**) “minimum node size”, using a random forest model with 1,000 trees. Bayesian optimisation is an informed search method where each iteration learns from the last. The optimal mtry and minimum node size is highlighted by orange. (**B**) mtry is the number of predictors (i.e., antigens) that will be randomly sampled for each split in the tree, with the default calculated as the square root of the number of *P. vivax* antigens. Increasing mtry improves the performance of the model yet decreases the diversity of the individual trees. Therefore, a balance between model performance and number of predictors in each tree is required. (**C**) minimum node size is the minimum number of data points in a node required for the node to be split further. In classification, the default is set to 1.

**Figure S2.**
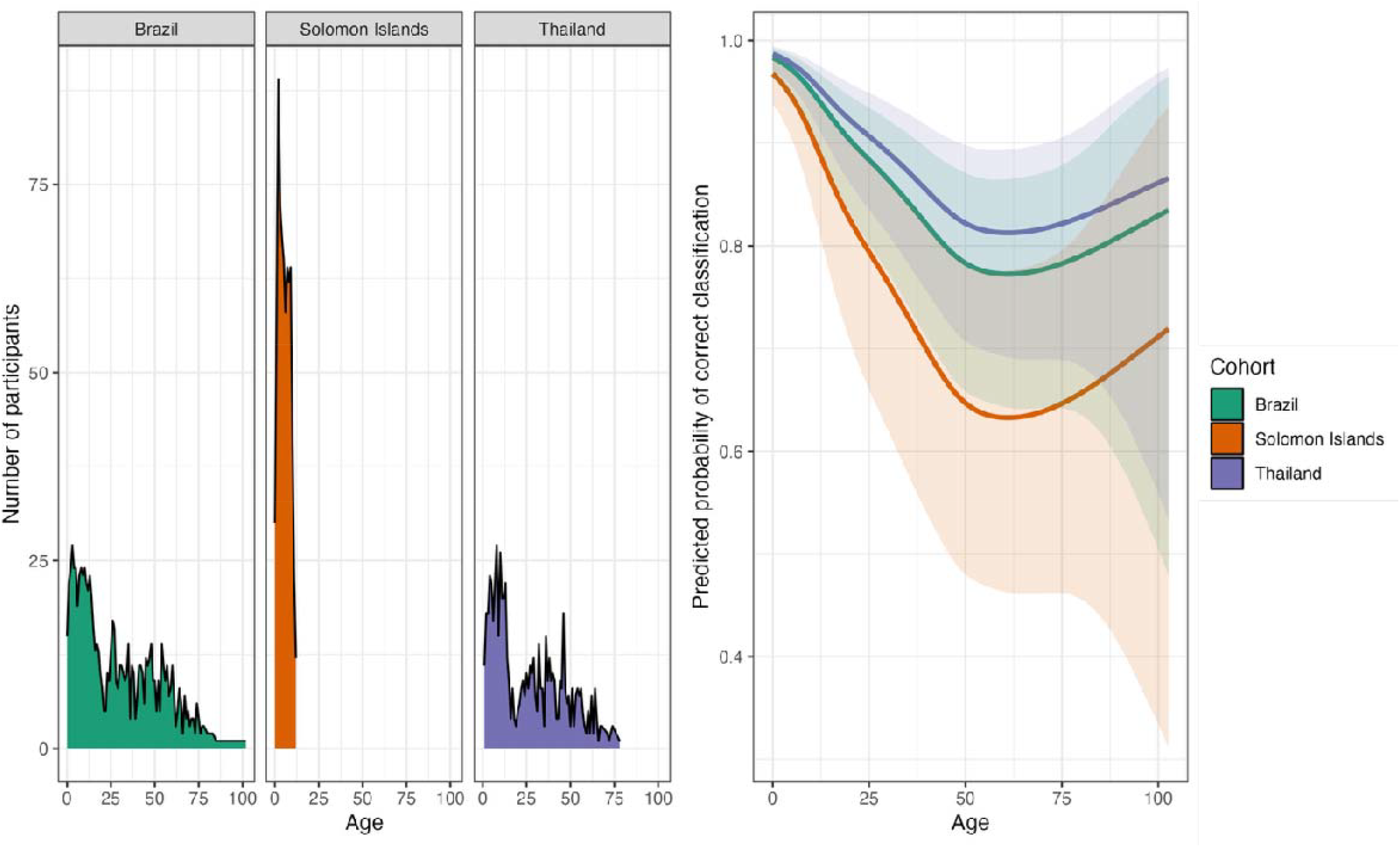
Relationship between age and random forest classification performance. (**A**) Age distribution of samples in each year-long longitudinal cohort (Brazil, green; Solomon Islands, orange; Thailand, purple). (**B**) Model-adjusted probability of a correct *P. vivax* classification by age for each cohort. Probabilities were estimated using a binomial generalised additive model with a penalised cubic regression spline for age. Correct *P. vivax* classification was defined as concordance between *P. vivax* serostatus and *P. vivax* qPCR results within the previous nine months (i.e., seropositive and qPCR positive, or seronegative and qPCR negative).

**Figure S3.**
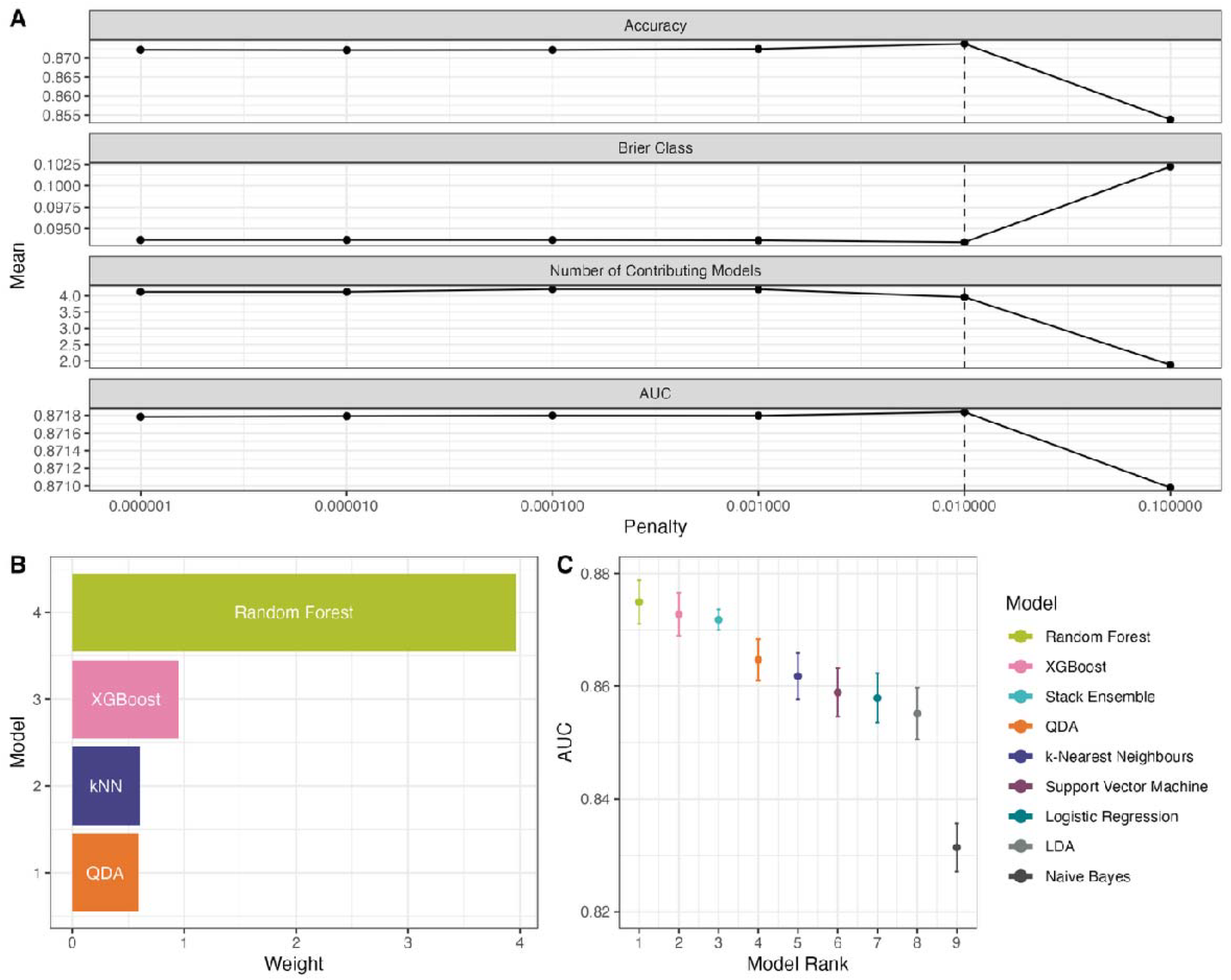
Evaluation of the multi-algorithm ensemble method (Stack Ensemble). (**A**) Mean performance metrics, including accuracy, Brier class, the number of contributing models and area under the receiver operator characteristic (AUC), across different LASSO regularisation penalties. The dashed line indicates the optimal penalty (0.001) that maximises performance across all four metrics. (**B**) Stacking coefficients for each of the four retained supervised machine learning classification models at the optimal penalty (0.001), where kNN refers to k-Nearest Neighbours. (**C**) Comparison of AUCs between the stack ensemble and the individual models shown in Figure 4A.

**Figure S4.**
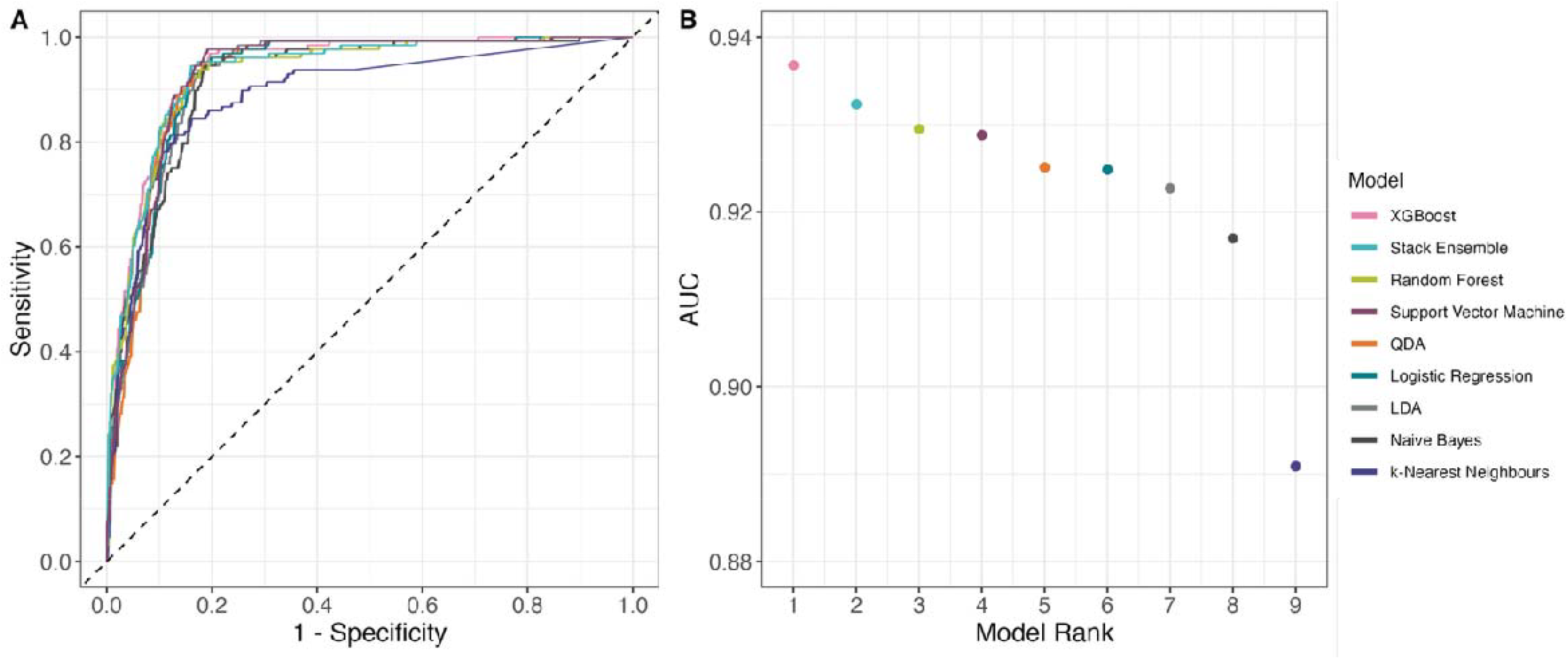
Validation of the multi-algorithm stacks ensemble using an external dataset from Indonesia^41^. (**A**) The receiver operator characteristic (ROC) curve for each model. (**B**) The area under the receiver operator characteristic (AUC) values ranked from highest to lowest for each unique supervised machine learning classification model and the stack ensemble.

**Figure S5.**
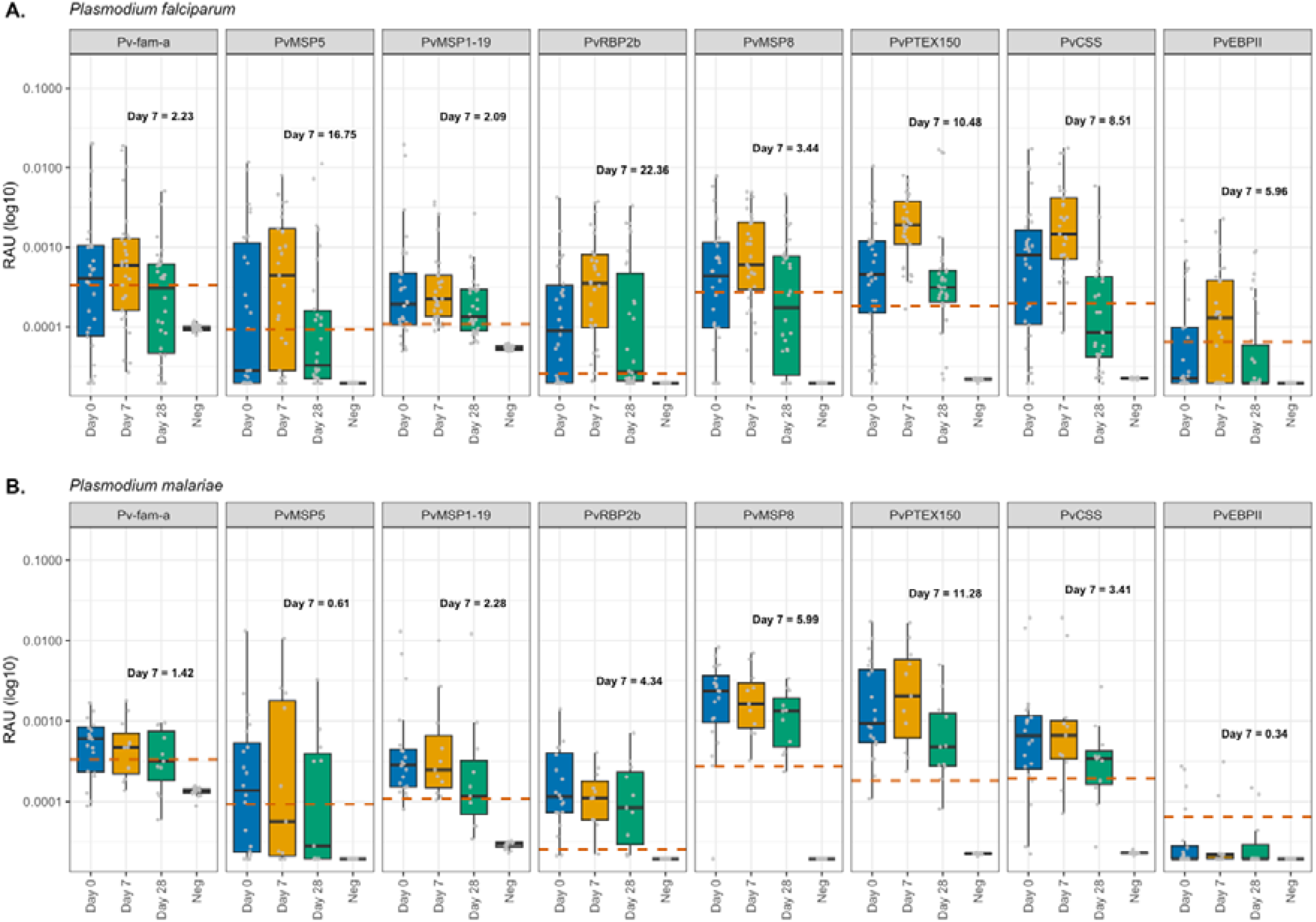
IgG antibody levels against 8 *Plasmodium vivax* proteins in patients following a clinical *P. falciparum* or *P. malariae* infection from Sabah, Malaysia. Antibody levels were measured in a single replicate against the top 8 *P. vivax* antigens using a multiplexed antibody assay and expressed as relative antibody units (RAU). Individual patients had longitudinal samples obtained and run at the time of diagnosis (day 0) with (**A**) *P. falciparum* or (**B**) *P. malariae*, and days 7 and 28 following enrolment in Sabah, Malaysia. (**A**) For *P. falciparum*, sample sizes were n = 30 at day 0, n = 29 at day 7, and n = 30 at day 28. (**B**) For *P. malariae*, sample sizes were n = 20 at day 0, n = 11 at day 7, and n = 11 at day 28. Negative samples in this figure indicate controls run at the same time of the experiment (*P. falciparum* n=19; *P. malariae* n=16). The dashed red lines indicate the seropositivity cut-off, defined as the mean RAU of the malaria-naïve negative-control samples (n=359) plus two standard deviations for each antigen. One isolate was excluded due to an outlier RAU value (>0.01) for PvCSS. The boxplots indicate the median, 25th and 75th percentiles with the whiskers showing the 2.5 and 97.5 percentiles, and dots representing outliers.

**Figure S6.**
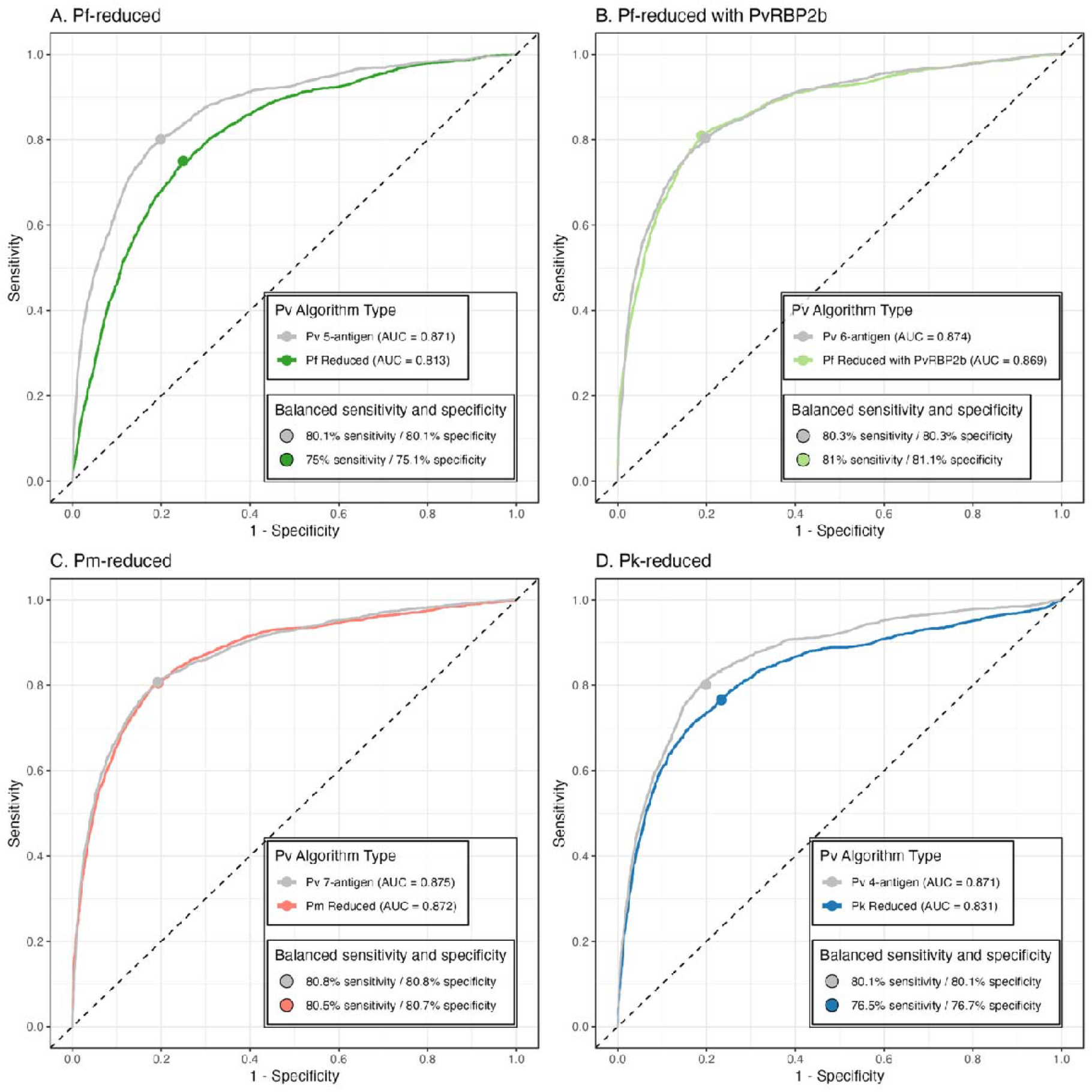
Comparison of the top performing *P. vivax* serological exposure classifiers minimising cross-reactivity with co-endemic *Plasmodium* species using a less conservative definition of cross-reactivity. Receiver operator characteristic (ROC) curves are shown for the top-performing PvSeroTAT antigen combination (grey) alongside antigen combinations defined to minimise reactivity with markers associated with *P. falciparum* (Pf, greens) and *P. malariae* (Pm, pink). A less conservative threshold of ≥10-fold-change above the seropositivity cut-off at day 7 was used to define antigens cross-reactive with *P. vivax* (Figure S5). (**A**) The Pf-reduced classifier includes PvMSP1-19, Pv-fam-a, PvCSS, PvEBPII and PvMSP8 (dark green), while the (**B**) Pf-reduced classifier with PvRBP2b additionally incorporates PvRBP2b (light green). (**C**) The Pm-reduced classifier includes PvMSP1-19, Pv-fam-a, PvMSP5, PvCSS, PvEBPII, PvMSP8 and PvRBP2b (pink) and (**D**) the Pk-reduced classifier includes PvMSP5, PvEBPII, PvPTEX150 and PvRBP2b (blue). The points on the ROC curve indicate the balanced sensitivity and specificity. The Area under the ROC curve (AUC) values are shown for each classifier. Each random forest classifier is run with 1,000 trees and default hyperparameters.

**Table S1.** Area Under the Receiver Operator Characteristic Curve (AUC) for each single-antigen classification model based on unique serological exposure markers (SEM).

| SEM | AUC | Standard Error | Rank |
| --- | --- | --- | --- |
| PvRBP2b | 0.832 | 0.012 | 1 |
| PvMSP1-19 | 0.786 | 0.013 | 2 |
| Pv-fam-a | 0.765 | 0.014 | 3 |
| PvEBPII | 0.755 | 0.014 | 4 |
| PvMSP8 | 0.740 | 0.014 | 5 |
| PvPTEX150 | 0.739 | 0.014 | 6 |
| PvMSP5 | 0.733 | 0.014 | 7 |
| PvCSS | 0.729 | 0.014 | 8 |
| PvRAMA | 0.690 | 0.015 | 9 |
| Pvs16 | 0.658 | 0.015 | 10 |

**Table S2.** Table of all the combinations for 2 to 8 antigens. (External Table).

**Table S3.** Adjusted odds ratios for a correct *P. vivax* serostatus classification estimated using a binomial generalised additive model with a logit link. Odds ratios are shown with 95% confidence intervals and are adjusted for *P. vivax* infection status, sex, cohort, and a penalised cubic regression spline for age. Female sex and current *P. vivax* infection status were used as reference categories. Negative controls were removed as they were all correctly classified in the random forest model and had incomplete age information.

| Contrast | Odds Ratio <sup>x</sup> | Standard Error | p-value |
| --- | --- | --- | --- |
| *P. vivax* Infection status |  |  |  |
| Current | Reference |  |  |
| Recent | 0.462 [0.215, 0.991] | 0.149 | 0.047 |
| Old | 0.121 [0.048, 0.304] | 0.047 | <0.001 |
| Never | 0.367 [0.181, 0.745] | 0.110 | 0.003 |
| Sex |  |  |  |
| Female | Reference |  |  |
| Male | 1.21 [0.978, 1.500] | 0.133 | 0.079 |
| Smooth Terms | Effective degrees of freedom | Chi squared test | p-value |
| Age | 5.156 | 146.4 | <0.001 |

<sup>x</sup> Data are presented as OR [95% confidence interval]

**Table S4.**
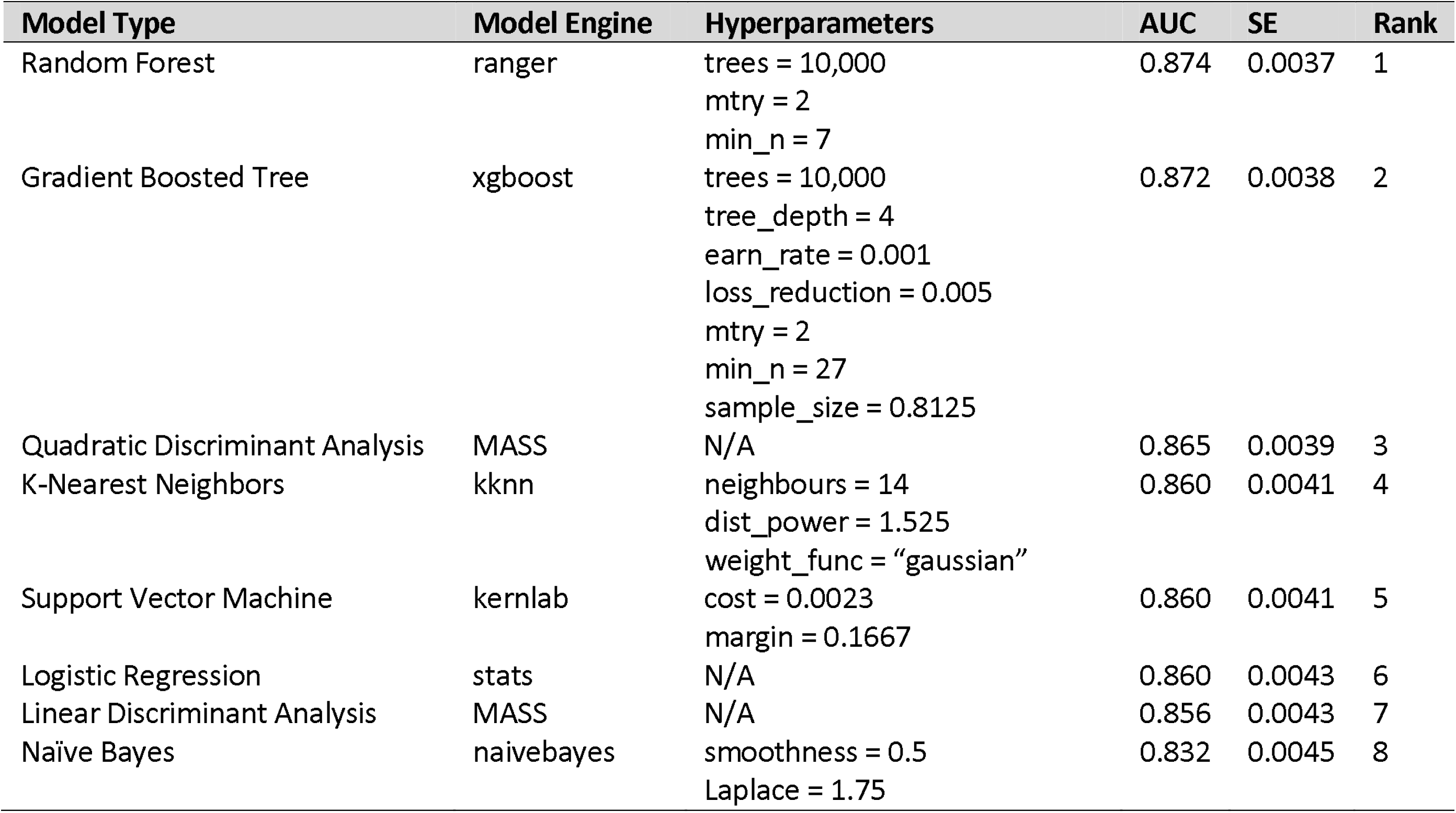
Model Comparison with optimised hyperparameters and Area Under the Receiver Operator Characteristic Curve (AUC) with standard error (SE), calculated using 10-fold cross-validation with five-repeats.

**Table S5.** Sequence comparison of the *P. vivax* proteins with their *Plasmodium* species orthologs.

| *P. vivax* |  | *P. falciparum* (3D7) |  |  |  |  |  |
| --- | --- | --- | --- | --- | --- | --- | --- |
| Gene ID (PlasmoDB) | Protein Name | Top Hit P | Syteny P | Identity (%) P | Similarity (%) P | Top Hit N | Identity (%) N |
| PVX_096995 | Pv-fam-a | PF3D7_0830500 | N | 20.7 | 35.5 | XP_001349226.1 | 32.08 |
| PVX_003770 | PvMSP5 | PF3D7_0206900 | Y | 22.7 | 32.6 | XP_002585389.1 | 46.54 |
| PVX_099980 | PvMSP1-19 | PF3D7_0930300 | Y | 37.4 | 55.1 | XP_001352170.1 | 46.73 |
| PVX_094255 | PvRBP2b | PF3D7_1335400 | N | 21.3 | 41.4 | Q8I4R2.1 | 23.72 |
| PVX_097625 | PvMSP8 | PF3D7_0502400 | Y | 36.3 | 53.5 | XP_001351583.1 | 47.20 |
| PVX_084720 | PvPTEX150 | PF3D7_1436300 | NA | 34.4 | 49.8 | XP_001348518.1 | 48.36 |
| PVX_086200 | PvCSS | PF3D7_1404700 | Y | 40.4 | 54.6 | XP_001348217.1 | 60.85 |
| KMZ83376.1<sup>a</sup> | PvEBP1I | NA | NA | NA | NA | 4K2U_A | 32.13 |
| *P. vivax* |  | *P. malariae* (UG01) |  |  |  |  |  |
| Gene ID (PlasmoDB) | Protein Name | Top Hit P | Syteny P | Identity (%) P | Similarity (%) P | Top Hit N | Identity (%) N |
| PVX_096995 | Pv-fam-a | PmUG01_02011300 | Y | 37.6 | 48.1 | SBT01095.1 | 61.42 |
| PVX_003770 | PvMSP5 | PmUG01_04025700 | Y | 31.8 | 43.6 | SBT70631.1 | 44.81 |
| PVX_099980 | PvMSP1-19 | PmUG01_07042000 | Y | 42.3 | 57.1 | ALP00323.1 | 56.48 |
| PVX_094255 | PvRBP2b | PmUG01_08058500 | N | 33.1 | 56.4 | SBS92940.1 | 32.90 |
| PVX_097625 | PvMSP8 | PmUG01_06023900 | Y | 56.8 | 74 | XP_028859893.1 | 59.53 |
| PVX_084720 | PvPTEX150 | PmUG01_13024000 | Y | 44.5 | 59.1 | SBT72076.1 | 43.65 |
| PVX_086200 | PvCSS | PmUG01_13053600 | Y | 51.6 | 65.6 | XP_028863921.1 | 61.94 |
| KMZ83376.1<sup>a</sup> | PvEBP1I | NA | NA | NA | NA | XP_028863493.1 | 43.79 |
| *P. vivax* |  | *P. knowlesi* (H) |  |  |  |  |  |
| Gene ID (PlasmoDB) | PlasmoDB | Top Hit P | Syteny P | Identity (%) P | Similarity (%) P | Top Hit N | Identity (%) N |
| PVX_096995 | Pv-fam-a | PKNH_0200900 | Y | 41.1 | 50.9 | OTN66147.1 | 70.28 |
| PVX_003770 | PvMSP5 | PKNH_0414200 | Y | 49.9 | 65.4 | AAT77929.1 | 46.26 |
| PVX_099980 | PvMSP1-19 | PKNH_0728900 | Y | 64.2 | 74.4 | AZL87433.1 | 81.31 |
| PVX_094255 | PvRBP2b | PKNH_0700200 | N | 29.1 | 52.2 | XP_038969555.1 | 28.81 |
| PVX_097625 | PvMSP8 | PKNH_1031500 | Y | 83.4 | 91.2 | AFL93300.1 | 84.52 |
| PVX_084720 | PvPTEX150 | PKNH_0422900 | Y | 73.2 | 83.9 | OTN66840.1 | 72.02 |
| PVX_086200 | PvCSS | PKNH_1353400 | Y | 69.0 | 77.7 | XP_038969555.1 | 69.53 |
| KMZ83376.1<sup>a</sup> | PvEBP1I | NA | NA | NA | NA | QPL17772.1 | 35 |

Full-length protein sequences were compared using the orthologs listed in PlasmoDB. For *P. falciparum*, the 3D7 strain was used; *P. malariae* the UG01 strain and *P. knowlesi* the H strain (which was consistent with A1H1 strain). Protein construct sequences were compared using NCBI BlastP (any strain) to identify the top hits. “P” denotes PlasmoDB pipeline and “N” denotes the NCBI pipeline. NA, no matches. Proteins are ordered by chromosome order in *P. vivax*.
<sup>a</sup> GeneBank ID.

**Table S6.** Calculation of the seropositivity cut-off based on malaria-naïve negative control cohorts (n=359) and fold-change in relative antibody units at day 7 per *P. vivax* antigen for patients following a clinical *P. falciparum* (n=30) or *P. malariae* (n=20) infection.

| Pv Antigen | Malaria-naïve negative control RAU<sup>1</sup> |  | Seropositivity cut-off<sub>2</sub> | Fold-change at day 7<sup>3</sup> |  | Fold-change at day 7<sup>4</sup> |
| --- | --- | --- | --- | --- | --- | --- |
|  | Mean | Sd |  | *Pf* | *Pm* | *Pk*<sup>4</sup> |
| Pv-fam-a | 0.000056 | 0.000139 | 0.000333 | 2.23 | 1.42 | 13.39 |
| PvMSP5 | 0.000022 | 0.000036 | 0.000093 | 16.75 | 0.61 | 4.70 |
| PvMSP1-19 | 0.000023 | 0.000043 | 0.000109 | 2.09 | 2.28 | 43.98 |
| PvRBP2b | 0.000020 | 0.000003 | 0.000026 | 22.36 | 4.34 | 3.00<sup>5</sup> |
| PvMSP8 | 0.000035 | 0.000118 | 0.000271 | 3.44 | 5.99 | 17.07 |
| PvPTX150 | 0.000055 | 0.000063 | 0.000182 | 10.48 | 11.28 | 6.08 |
| PvCSS | 0.000040 | 0.000077 | 0.000194 | 8.51 | 3.41 | - |
| PvEBPII | 0.000021 | 0.000021 | 0.000064 | 5.96 | 0.34 | 1.33 |

Pv: *P. vivax*; Pf: *P. falciparum*; Pm: *P. malariae*;
<sup>1</sup> Calculated from malaria-naïve negative controls ( $N=359$ ); $n=1$ was removed with very high RAU > 0.01 for PvCSS.
<sup>2</sup> Calculated as the mean plus two standard deviations.
<sup>3</sup> Calculated from the clinical *P. falciparum* ( $n=30$ ) and *P. malariae* ( $n=20$ ) infections from Sabah, Malaysia.
<sup>4</sup> Values from Longley et al. 2022 *Cell Reports Medicine* Table 4. There was no value for PvCSS.
<sup>5</sup> PvRBP2b value taken from PvRBP2b<sub>161-1454</sub>.

